## Supplementary Document 1 for "A randomised survey of the quality of antibiotics and other essential medicines in Indonesia, with volume-adjusted estimates of the prevalence of substandard medicines"

### Detailed description of methods used in the STARmeds study of medicine quality

Elizabeth Pisani, on behalf of the STARmeds Study Team.

**DATA SOURCES** 3

**SELECTION OF MEDICINES** 5

**SAMPLING** 7

**SELECTION OF DISTRICTS** 7
**SELECTION OF OUTLET TYPES** 9
**SAMPLE SIZE CALCULATION** 10
**SAMPLE FRAME CONSTRUCTION** 11
GREATER JAKARTA 12
SELECTION OF PHARMACIES AND MEDICINE SHOPS 13
SELECTION OF OTHER SOURCES OF MEDICINE, OFFLINE 13
ONLINE SAMPLING FRAME 13
ALLOCATION OF MEDICINES ACROSS OUTLETS 14

**SAMPLE COLLECTION** 14

**DATES AND LOGISTICS** 14
**FIELDWORKER TRAINING AND SUPERVISION** 15
**DATA COLLECTION TOOLS** 15
**SAMPLE COLLECTION PROCEDURES** 16
ONLINE SAMPLES 17
**SAMPLE PROCESSING AND STORAGE** 18
SAMPLE INTAKE AND PACKAGING INSPECTION 18
ADDITIONAL DATA ENTRY 18
VERIFICATION AND STORAGE 18
**SAMPLE TRIAGE** 18
**LABORATORY TESTING** 19
APPEARANCE 21
SAMPLE PREPARATION 21
IDENTIFICATION 22
ASSAY TESTING 22
DISSOLUTION TESTING 22
IMPURITY TESTING 23

**VERIFICATION OF AUTHENTICITY** 24

**QUALITY DEFINITIONS** 24

SUBSTANDARD VERSUS FALSIFIED MEDICINES 26

**DATA MANAGEMENT** 26

ETHICS AND REPORTING 26

ETHICAL AND POLICY REVIEW 26
HEALTH PROTOCOLS 27
MEDQUARG 27

REPORTING RAW PREVALENCE OF POOR QUALITY MEDICINES 27

METHODS FOR DEVELOPING NATIONAL LEVEL ESTIMATES 28

CALCULATING PREVALENCE BY PRODUCT 28
CALCULATING MARKET VOLUME BY PRODUCT ERROR! BOOKMARK NOT DEFINED.

WEIGHTED ESTIMATES ERROR! BOOKMARK NOT DEFINED.

REFERENCES 33

### Data sources

Tables A1 and A2 list the data sources referred to in the remainder of this document, and in associated papers.

**Table A1: Data sources, medicines**

| | Dataset name and status | Description | MA-holder | API | Dose, form | Brand | MA number | Sales volume | Sales value | Price* | MRP | Quality tests |
|---|---|---|---|---|---|---|---|---|---|---|---|---|
| 1 | BPOM registration (Open access) | List of all medicines with current valid market authorisation in Indonesia, 2020-2022 | Y | Y | Y | Y | Y | N | N | N | N | N |
| 2 | e-catalog (Requested) | Winners of annual public procurement tender, per province, all medicines, 2018-2022 | Y | Y | Y | Y | Y | Y | Y | Y | N | N |
| 3 | IQVIA market (Paid) | Brands recorded in audit of 250 hospitals, 1064 pharmacies and 250 non-prescription medicine shops; 10 APIs, 2020-2022 | Y | Y | Y | Y | N | Y | Y | Y | N | N |
| 4 | STARmeds (Anonymised data shared) | Medicines sampled in survey in Greater Jakarta, 6 other districts and online, 5 APIs, 2022 | Y | Y | Y | Y | Y | N | N | Y | Y | Y |
| 5 | BPOM quality history (Requested) | Confidential listing of medicines testing substandard or confirmed falsified in post-market surveillance | Y | Y | Y | Y | Y | N | N | N | N | Y |

*The e-catalog price is the winning price per province. The IQVIA price is the manufacturer's published list price, exclusive of VAT. The STARmeds price is the price actually paid for the sample by sample collectors. For overtly sampled products which were free to patients, we recorded the facility's cost price.

BPOM: Indonesian medicine regulator
MA: Market Authorisation.
MRP: Maximum retail price (In Indonesian this is the Harga Eceran Tertinggi, or HET)
API: Active Pharmaceutical Ingredient

**Table A2: Data sources, other**

| | Dataset | Source and description | Data/indicators used |
|---|---|---|---|
| 6 | Insurance utilisation data | Sample or summary data published by BPJS Kesehatan, the national health insurer that reports covering 80% of patients | Ranking of conditions by prevalence<br>Ranking of conditions by spending<br>Ranking of reimbursed medicines by spending<br>Total value of claims per 1,000 insured. |
| 7 | Essential medicines list | List of medicines that should be available free to all insured patients, by facility level, MoH | Medicines considered essential at primary level |
| 8 | Basic Health Indicators | Regular national survey of health indicators, from representative survey of households and primary health centres, from BPS and MoH | Prevalence of common diseases |
| 9 | Census | 2020 national census from BPS. All data analysed by district. | Population<br>Population density |
| 10 | SUSENAS | Biannual representative survey of over a million people; BPS. All data analysed by district, for 2018 | Per capita income by district<br>% of population reporting health insurance, by source of funding<br>Health spending per capita |
| 11 | Village Census | A periodic census of facilities throughout Indonesia, at the level of over 84,000 villages; BPS. 2018 data. | Pharmacies per 100,000 population<br>Hospitals per 100,000 population |
| 12 | Google | Dataset generated by analysing online search strategies using Indonesian variants on "buy medicine", "cheap medicine", and lists of medicine names. | Relative frequency of online searches, preceding 6 months |

BPJS Kesehatan: Indonesia's national health insurance agency
MoH: Indonesian Ministry of Health
BPS: Indonesia's national statistics agency

#### Selection of medicines

In the following description, the numbers in square brackets refer to the numbered data sources in Tables A1 and A2. Where these are publicly available, references are given.

We selected medicines for inclusion based primarily on their public health importance, considering:

- Presence on the national essential medicines list; [Table A1/A2 Data source 7]
- Volumes sold through public procurement and private retail outlets, [Data sources 2, 3];
- Ranking of medicine in National Insurer's list of reimbursed medicines, [6]
- Burden of diseases/conditions treated. [6, 8]

We aimed to select medicines for conditions across a variety of therapeutic categories, treating high-burden conditions.

We also took feasibility into account. Medicines had to be within our budget to acquire and test [3, laboratory tariff schedule], available in retail pharmacies, feasible to acquire using mystery shoppers, and not requiring cold chain handling. We considered product variation, favouring medicines with a higher number of authorised sellers. [1] We excluded medicines for vertical health programmes which are procured partly through parallel supply chains (for example HIV and TB medicines).

In addition, because we wished purposively to include medicines at potential risk for falsification in the study, we considered history of falsification or out-of-specification testing over the preceding three years [5], and used Google Analytics to investigate the frequency of different medicine names featuring in Google searches by Indonesian users over the preceding 6 months [12]. Most were classified as psychoactive medicines or were 'lifestyle' medicines for erectile dysfunction or dieting. However, two medicines which featured highly in Google searches, and which had other potential risks for falsification, also fit our criteria for public health importance. There were allopurinol (a medicine widely used to control gout; allopurinol has been growing in popularity among non-medical users of street drugs in Jakarta and was seized in a police raid of medicines made by falsifiers in the Greater Jakarta region in January 2022) and cefixime (a relatively expensive antibiotic which is frequently prescribed at primary level, but not covered by insurance in public primary facilities, and which featured on the regulator's list of falsified medicines identified between 2018-2020).

Based on these criteria, we drew up a short-list of 12 medicines. All are on the national essential medicines list, although cefixime is only recommended at the referral level, not for primary care. Table A3 provides details of the key characteristics of the short-listed medicines.

**Table A3: Characteristics of short-listed medicines**

| No. | Medicine | Therapeutic use | Ranking within ATC | | IQVIA 2019 [3] | | Number of authorization holders 2021 [1] | Feasibility |
|---|---|---|---|---|---|---|---|---|
| | | | Public procurement volume, 2018 [2] | Out of specification, 2016-19* [5] | Volume, all formulations | Median price, highest-volume formulation | | |
| 1 | Albendazole | Anthelmintic | 1st ATC P | 1st ATC P | N/A | N/A | 3 | High |
| 2 | Allopurinol | Enzyme inhibitor | 1st ATC M | 4th, ATC M; falsified | 261,560,080 | 234 (100mg tablet)<br>777 (300mg tablet) | 50 | High |
| 3 | Amlodipine | Anti-hypertensive | 1st ATC C | 4th ATC C | 524,451,810 | 1,516 (5mg tablet) | 74 | High |
| 4 | Amoxicillin | Antibiotic | 1st ATC J | 1st ATC J | 431,548,142 | 588 (500mg tablet)<br>1473 (500mg capsule)<br>503 (125mg/5 ml dry syrup) | 58 | Medium |
| 5 | Cefixime | Antibiotic | 8th ATC J | 6th ATC J; falsified | 146,177,158 | 11,589 (100mg capsule) | 42 | Medium |
| 6 | Cotrimoxazole | Antibiotic | 3rd Combination medicine | 1st Combination medicine | 56,828,900 | 263 (480mg tablet) | 49 | Medium |
| 7 | Dexamethasone | Corticosteroid | 1st ATC H | 1st ATC H | 577,110,925 | 90 (0.5mg tablet) | 44 | High |
| 8 | Glibenclamide | Anticholesterol | 11th ATC A | 2nd ATC A | 116,295,550 | 159 (5mg tablet) | 11 | High |
| 9 | Haloperidol | Anti-psychotic | 2nd ATC N | 1st ATC N | 21,508,638 | 235 (5mg tablet) | 9 | Low |
| 10 | Paracetamol | Analgesic | 1st over the counter | 1st over the counter | 1,654,542,677 | 123 (500 mg tablet)<br>358 (120mg/5 ml syrup) | 102 | Low |
| 11 | Prednisone | Corticosteroid | 2nd ATC H | 2nd ATC H; falsified | 203,581,000 | 55 (5mg tablet) | 21 | High |
| 12 | Trihexyphenidyl | Antispasmodic | 1st ATC N | 1st all falsified medicines | N/A | N/A | 5 | Low |

ATC: Anatomical Therapeutics Category. Information on the categories can be found at https://www.whocc.no/atc_ddd_index/
The out of specification data refers to samples collected in post-market surveillance conducted by the Indonesian regulator BPOM that failed visual inspection or assay or dissolution testing. The falsification data are confirmed falsified samples identified by the regulator between 2018 and 2020, with the exception of allopurinol, which was reported by police in 2022.

The shortlist was discussed by a national technical working group on medicine quality, including members from the Ministry of Health, the National Health Insurer, the National Medicine Regulator, health professional associations and academia. Paracetamol was rejected because drawing up a sample frame for an over-the-counter medicine widely available in supermarkets, corner shops and roadside kiosks would be too challenging. Psychoactive trihexyphenidyl and haloperidol had been under recent strict oversight by the narcotics control board, and was considered impractical to acquire through mystery shopping. Although anthelmintic albendazole is the most commonly used in its anatomical-therapeutic class, there are only three authorised products on the Indonesian market, limiting the likely variety of price or quality. Brand variation was similarly limited for glibenclamide.

Of the shortlisted medicines, the group recommended including the highest-volume antibiotic, corticosteroid, and chronic disease medicines (amoxicillin, dexamethasone and amlodipine respectively), and the two non-psychoactive medicines considered at risk for falsification (allopurinol and cefixime). Since dry syrup formulations of amoxicillin are very commonly prescribed for paediatric use, it was recommended that this formulation be included as well as the 500mg tablet formulation. The working group did not recommend making a distinction between tablets, capsules and caplets of the chosen dosages for other medicines (though practically, only the antibiotics are available in capsule form in the Indonesian market).

The selected medicines were:

- Allopurinol (100mg tablets; 300mg tablets)
- Amlodipine (5mg tablets)
- Amoxicillin (500mg tablets or capsules; 125 mg/ml dry syrup)
- Cefixime (100mg tablets or capsules)
- Dexamethasone (0.5mg tablets)

By regulation, all of the selected medicines should be dispensed only against prescription in Indonesia.

### Sampling

#### *Selection of districts*

The selection of study districts was largely purposeful. The total number (8) was chosen to achieve a balance between diversity, explanatory power, feasibility and budget. We listed demographic indicators using data from the 2020 census [9] and district-level health indicators from national surveys.[8, 10, 11] We chose three provinces which reflected Indonesia's geographic diversity -- one each from Western, Central and Eastern Indonesia. Within those provinces, we chose the provincial capital and one or two candidate rural districts. In addition, we used probability proportional to size to randomly select two districts in Greater Jakarta, as described in the section on sample frame construction. Greater Jakarta is a metropolitan agglomeration of close to 30 million people, which includes six districts in the capital, Jakarta, and eight districts in neighbouring West Java and Banten provinces.

In selecting districts outside of Greater Jakarta, we took feasibility and potential utility into account; we preferred districts where team members had existing links with researchers or health policy actors interested in medicine pricing and quality, or where comparable data were available. We excluded districts that were more than 12 hours' journey from their provincial capital.

**Table A4: Characteristics of study areas and districts**

| | East Jakarta | Bekasi City | Surabaya | Malang District | Medan | Labuhan Batu | Kupang City | TTS | National median (of 514 districts) |
|---|---|---|---|---|---|---|---|---|---|
| Population (millions) [9] | 2.9 | 2.9 | 2.9 | 2.6 | 2.6 | 0.48 | 0.42 | 0.47 | 0.27 |
| Population per km$^2$ [9] | 15,480 | 14,103 | 8,225 | 733 | 8,525 | 225 | 2,335 | 118 | NA |
| % reporting JKN [10] | 79.2 | 70.9 | 51.4 | 39.0 | 60.7 | 48.6 | 67.7 | 56.5 | 61.4 |
| % reporting subsidized JKN [10] | 45.9 | 17.7 | 17.5 | 20.4 | 26.5 | 21.1 | 27.9 | 49 | 41.8 |
| Number of pharmacies [11] | 258 | 322 | 552 | 179 | 396 | 61 | 76 | 10 | 26 |
| Pharmacy per 100,000 population | 8.9 | 11.1 | 19.2 | 6.9 | 17.5 | 12.6 | 18.1 | 2.2 | 8.9 |
| Number of hospitals [11] | 42 | 51 | 54 | 23 | 95 | 11 | 11 | 2 | 3 |
| Hospital per 100,000 population | 1.4 | 1.8 | 1.9 | 0.9 | 4.2 | 2.3 | 2.6 | 0.4 | 1.0 |
| Self-reported health spending per capita, IDR/year [10] | 98,333 | 146,667 | 134,500 | 105,800 | 50,000 | 49,250 | 40,000 | 34,000 | 50,000 |
| Amount paid for JKN claims/ per member [Million IDR/year] [6] | 866.4 | 635.6 | 929,9 | 321.3 | 734.0 | 312.0 | 1,012. | 26.8 | 189.0 |

JKN: *Jaminan Kesehatan Nasional* -- National Health Insurance
IDR: Indonesian rupiah. Average exchange rate for year of data collection: USD 1 = IDR 14,870

Proposed districts were discussed with a national technical working group on medicine quality. The group proposed one change, substituting Labuhan Batu in North Sumatra for a previously suggested inland district because it is a port area with frequent cross-straits trade with Malaysia and thus was thought to be a potential entry point for unregistered medicines. In each selected district, we contacted the local government, provided them with the study protocol and other information, and obtained written permission to undertake the study.

Table A4 shows the districts selected, with key characteristics. Data sources [in square brackets] are listed in Tables A1 and A2.

#### Selection of outlet types

We wished to get as comprehensive a picture of medicine quality across the Indonesian market as possible. When sampling prescription medicines for post-market surveillance, the national medicine regulator BPOM samples from registered pharmacies, public and private hospitals, and primary health clinics. After consultation with BPOM and the Ministry of Health, we included all of these outlet types, as well as some not included in routine post-market surveillance because they are not technically permitted to dispense prescription medicines. These were: over-the-counter medicine shops; doctors and midwives in private practice; and general internet marketplaces. (In very remote areas, prescription medicines are also occasionally sold at roadside kiosks (*warung*), and by itinerant vendors in weekly markets, but listing and sampling these randomly was not feasible.) We also included markets specialising in sales of bulk medicines. These exist only in Greater Jakarta, and are believed to sell mostly to health care providers and informal medicines sellers, especially from remote areas. The regulatory status of individual sellers in the market at the time of sampling was fuzzy. BPOM considers them unregulated and did not sample from them at the time of our study, but they are tolerated by the district and provincial government.

##### Online outlets

At the time of sampling, the Indonesian Ministry of Health had certified four pharmacy or aggregator applications for smartphones. These use geolocation to find a registered pharmacy near the patient, and to organise medicine delivery. They operate in large cities. A further three companies were registered to sell prescription medicines online through centralised apps or websites.

Technically, these were the only companies allowed to sell prescription medicines online at the time of the survey. However, the Ministry of Health website recommended various other sites for online purchases during COVID-related restrictions on movement. In addition, Indonesia's largest online general marketplace Tokopedia -- equivalent to Alibaba or Amazon in other markets -- operated a verification scheme which provided "official store" badges to licensed bricks-and-mortar pharmacies who wished to offer prescription medicines online. A Google search for "buy medicine online" and similar search terms (in Indonesian: "*beli obat online", "beli amlodipin online"* etc.) revealed that unlicensed individual sellers also sold medicines on Tokopedia, other general marketplaces such as BliBli and Lazada, through unlicensed medicine-specific outlets such as HDMall, and through social media sites including Facebook, Instagram and Twitter.

We grouped all identified online sampling outlets into four categories, which we classified as shown in Table A5.

**Table A5: Classification of on-line sources of medicine**

| Definition of outlet type | Legal status | STARmeds classification; notes |
|---|---|---|
| Registered apps using geo-location | Regulated | Regulated; available in major cities. n=4 |
| Registered on-line pharmacies | Regulated | Regulated; n=3 |
| Licensed bricks-and-mortar pharmacies selling online as official stores on the Tokopedia platform | Unregulated | Semi-regulated |
| Individual or unlicensed sellers on Tokopedia and other general goods marketplaces; unregistered online medicine websites; social media sellers | Unregulated | Unregulated |

We noted the position of unregulated sellers in the page rankings of search-results for medicines (searched for by medicine name or brand), as well as the volumes sold. (For example, a search for amlodipine might yield 11,800 results on Tokopedia, the largest sales platform, with the first unregulated seller appearing on page 87 of 118; their sales figures might show that that they had previously made two sales of the medicine in question, compared with hundreds or thousands of units sold by verified sellers). These data were used to estimate online volumes, as described in the estimation section.

#### *Sample size calculation*

Accurate sample size calculation was complicated by two primary factors.
1) At the time of initial study design (when seeking funding and thus proposing a budget), the only information available on the background prevalence of substandard or falsified medicines in Indonesia was an annual performance indicator from the medicine regulator.

2) Since our intention was to use mystery shoppers, it was not possible to randomise at the level of the medicine, only at the level of the outlet. The assumption of a normal distribution which underlies traditional calculations of sample sizes for measurement of a proportion, or for the difference between two proportions, therefore does not hold, unless you also assume that poor quality medicines are randomly distributed between outlets. Previous research suggest that is very unlikely in the Indonesian market.(Hasnida, Kok, and Pisani 2021; Pisani et al. 2022)

WHO guidelines for conducting medicine quality surveys provide no pointers on sample size calculations in these circumstances.(WHO Expert Committee on Specifications for Pharmaceutical Preparations 2016) Few peer-reviewed studies of medicine quality report their sample size calculations. Those that do either differ in sampling design from the current study,(Schiavetti et al. 2018; Schäfermann et al. 2018) or assume quality is randomly distributed.(Khurelbat et al. 2014)

Since we had no way of estimating existing differences in the very prevalences we were seeking to measure, we assumed normal distribution and used the formula for sample size calculation for a proportion $n= p(1-p) \times (1.96*e)^2$ to estimate the number needed to estimate the prevalence of out-of-specification medicines within any subgroup (for example, by medicine, or by geographic region). Setting the desired confidence interval at 95% and using the WHO's blanket estimate of prevalence of poor quality medicines in middle income countries of 10% as guesstimate of prevalence,(World Health Organization 2017) that gave a minimum sample size of $(0.1x0.9)x(1.96*0.05)^{2,}$ or 138 for each sub-group.

Our maximum sample size was determined by budget. This set an initial ceiling of 1200 samples. We distributed these across medicines, districts and outlet types. The aim was to ensure where possible a minimum of 138 samples per medicine, and per district (across all medicines). This was not feasible in the case of amoxicillin dry syrup, because the large number of bottles required per sample limited the outlets we could approach for this medicine. In the smaller and less populous rural districts, it was also not feasible to assign such a large sample. On the advice of BPS statisticians, we stratified the sample distribution by number of retail outlets in each sampling area, as show in Table A6. We then assigned a weight to each of the strata to distribute the total sample size.

**Table A6: Sample distribution at study design phase, physical outlets and geolocation apps**

| Sampling area | Characteristic | Pharmacies listed by MoH | Stratum | Assigned sample |
|---|---|---|---|---|
| Greater Jakarta | Megacity | 1204 | 4 | 239 |
| Surabaya | Large city | 507 | 3 | 179 |
| Medan | Large city | 439 | 3 | 179 |
| Kupang | Small city | 84 | 2 | 119 |
| Malang district | Semi-rural | 213 | 2 | 119 |
| Labuhan Batu | Remote rural | 24 | 1* | 60 |
| Timor Tengah Selatan | Remote rural | 16 | 1* | 60 |

*No samples assigned for purchase from smartphone applications using geolocation

After we completed fieldwork, BPOM requested that we add to our power to detect differences between quality in the regulated and unregulated supply chain, including unregulated online sellers and the bulk medicine markets in Greater Jakarta. In response, we added a target of 70 additional samples from these markets.

#### *Sample frame construction*

Our sampling strategy was developed in consultation with experts from the sampling division of the Indonesian national statistics bureau, BPS. We adapted the approach, illustrated in Figure A1, according to the number of outlets and populations of the selected districts.

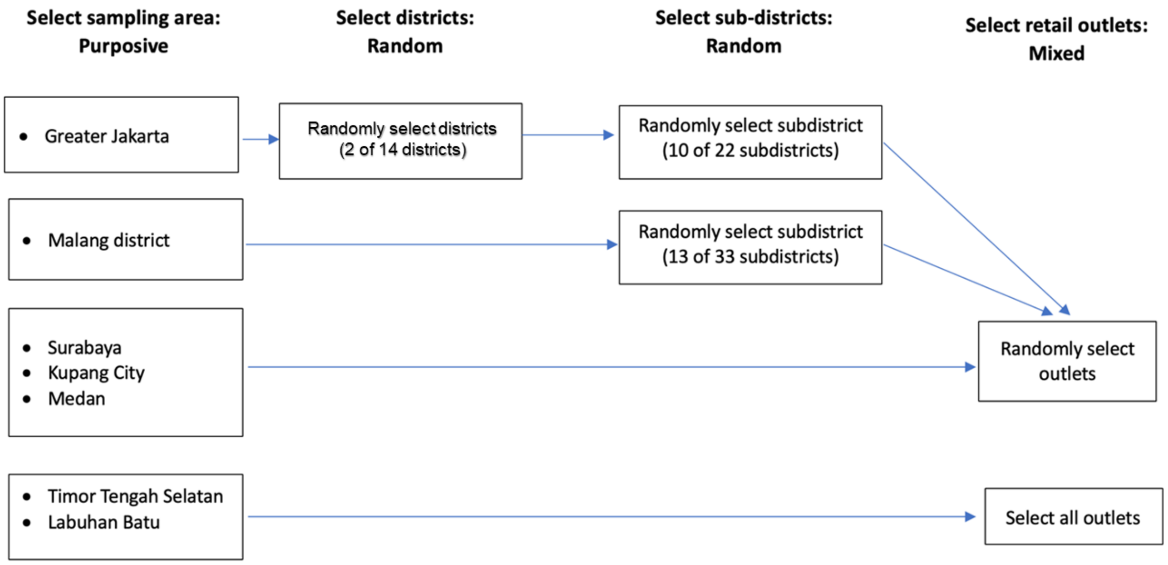

**Figure A1: Schematic illustration of sample frame construction**

- In Greater Jakarta, we used two-stage cluster sampling, selecting districts and sub-districts with probability proportional to size (PPS); then sampled outlets randomly;
- In Malang district, we used one-stage PPS sampling to select subdistricts; then sampled outlets randomly;
- In Surabaya, Medan and Kupang, we used simple random sampling for outlets;
- In Labuhan Batu and Timor Tengah Selatan (TTS) we used a take-all approach for pharmacies, and simple random sampling for other outlets.

#### Greater Jakarta

Since millions of people travel daily between numerous districts in the Greater Jakarta area, we treated it as a single sampling unit, and used two-stage randomised selection with probability of selection proportionate to the number of retail medicine outlets.

##### Districts

We first listed each of the 14 districts of Greater Jakarta, ordering them by the absolute number of pharmacies recorded in the district by the Village Census. [11] After consulting with staff at the Bureau of Statistics and ascertaining that the census data did not include individual outlets in well-known bulk medicine markets, we physically mapped shops within these markets, and added those outlets to the total count.

We chose two of the districts randomly, using the probability-proportional-to-size methodology with a target sample size of 100 pharmacies in two clusters.(Bierrenbach 2015). While it would have been possible for both selected districts to be in a single province, the final randomly selected districts were East Jakarta (in Jakarta province -- population 2.91 million) and Kota Bekasi (in West Java province, population also 2.91 million).

##### Sub-districts

To increase feasibility of field work in these large and populous districts, we further restricted sampling areas to sub-districts, also selected using probability proportionate to size. For the two selected districts, we collected listings or other data on retail pharmacies and medicine shops from additional sources to increase the accuracy of listing provided by MoH listings alone. These included:

- The district Department of Health
- The Indonesian Pharmacists' Association
- Online health services listings sites (e.g. goalkes.com)
- Online mapping applications (e.g. Google Maps, Waze)

We triangulated data across these sources, removed duplicates, summed the number of outlets by sub-district, and repeated the PPS exercise. We now aimed to distribute the 100 pharmacies across 10/22 clusters (sub-districts), with a target of 10 pharmacies per cluster.

Again, since we treated Great Jakarta as a single sampling unit, there was a possibility that all clusters would be selected from the same district. Randomisation in fact yielded a sample frame that included 5 subdistricts in each of the two districts.

In Malang district, we listed all sub-districts with the number of pharmacies in each district, and selected 13 of 33 sub-districts with probability proportionate to size.

#### Selection of pharmacies and medicine shops

##### Verification

For the selected subdistricts in Greater Jakarta, and for Surabaya city, we attempted to verify the continuing existence of every listed pharmacy by phone and through rapid field surveys. In all other districts, fieldworkers conducted full physical verification prior to outlet selection, providing photos and geo-locations for all verified pharmacies.

##### Random selection

For the chosen districts or subdistricts, we entered all verified pharmacies and medicine shops (including those in wholesale markets) into a database. We used Stata 17 to randomly select the target number of outlets in each area ("core outlets"). We also randomly selected alternate outlets (up to a third of the target number) in case sample collectors were unable to buy the designated medicine at the core outlet (for example because of a stockout). These were held in reserve by study staff, and communicated to sample collectors during data collection if necessary.

At the request of the medicine regulator, we collected additional samples from Greater Jakarta's bulk medicine markets at the end of the sampling period. (We had already collected 33 samples from these markets according to the initial sampling plan). We selected outlets for this additional sampling by simple random sampling from a verified listing of all individual outlets in these markets.

Because there were very few pharmacies in the two remote rural districts, no selection was made in those areas -- all were included in the sample frame.

Downloadable and reusable examples of the code used to generate the sample frame are available in our fieldwork Toolkit at: https://doi.org/10.7910/DVN/OBIDHJ

#### Selection of other sources of medicine, offline

We obtained lists of public and private health facilities from district health authorities. They also provided listings of doctors and midwives licensed for private practice but these were in some cases quite outdated. We triangulated with other data sources as available, including listings from professional associations and internet directories of health providers.

We entered the listings into a database by sampling area. We selected a district public hospital (in most districts there is only one), one private hospital, and one primary health centre (*puskesmas*) in each sampling area, using Stata 17's random generation function. We also randomly selected one alternate private hospital and health centre.

We selected doctors and midwives in private practice in the same way, randomly choosing five in each location, with three alternates.

#### Online sampling frame

In Greater Jakarta, Surabaya, Medan and Malang district, each data collector was assigned one of the four instant-delivery (geolocation-based) sales platforms in their individual sampling plan, at random. This accounted for 37 samples in all. Instant-delivery platforms were not available in other locations.

We listed all other identified sources of medicines online. Because we were unable to estimate the "size" of each cluster (the number of individual sellers on each platform) we purposively assigned buyers to target every one of the identified platforms.

##### Allocation of medicines across outlets

Because we wished to investigate the relationship between price and quality, we aimed to sample low cost and higher-cost versions of the same medicines from the same outlets.

At hospitals and from doctors and midwives, we did not use mystery shoppers, but openly requested to buy samples for the purposes of the study. In these settings, we aimed to sample a branded and an unbranded version of each of the study medicines. Branded medicines are rarely available at primary health centres, where we collected any available version of study medicines.

For retail outlets and the internet, medicines were generally distributed across outlets in clinically plausible pairs. Each outlet was assigned the same pair of medicines twice, one shopper targeting cheap versions and the other more expensive versions. We translated the targets into individual sample frames for each mystery shopper, grouped by sub-district or area. Each shopper had a single price point (cheaper or more expensive) across the whole sampling period, and was given a daily target, by medicine and outlet. Shoppers targeting the same outlet were assigned that location on different days.

### Sample collection

#### *Dates and Logistics*

We sampled in waves, by province, following the timetable shown in Table A7

**Table A7: Dates of sample collection**

| District | Training | Sampling and data entry |
|---|---|---|
| Greater Jakarta | 14 Feb 2022 | 15-20 Feb 2022 |
| Surabaya, Malang | 28 Feb 2022 | 1-5 Mar 2022 |
| Medan, Labuhan Batu | 21 Mar 2022 | 22-26 Mar 2022 |
| TTS | 28 Mar 2022 | 29 Mar – 2 Apr 2022 |
| Kupang | 4 Apr 2022 | 5 – 8 Apr 2022 |
| Online and bulk markets | 17-18 Jan 2022 | 15 Feb – 17 May 2022 |

We rented a house short-term for use a study hub in each district except Kupang, where we used a hotel. Each was equipped with air-conditioning, light boxes and dedicated tablets for sample photography and data entry. The study hub was used for sample processing, including additional data entry and sample storage, as well as STARmeds staff accommodation.

At each study hub, we had core study staff (mostly full-time STARmeds researchers) performing the following roles:

- Site supervisor: overall running of the site; sample intake; visual inspection; support for data entry staff;
- Fieldwork support: full-time phone support for sampling staff; troubleshooting of sample frame (e.g. providing alternate randomly-selected outlets in case of closure, flooding or stockouts); real-time monitoring of sampling progress;

- Logistics and finance manager: ensuring adequate supplies and connectivity; logging daily spending and collecting receipts; reimbursement of field workers; sample storage and shipment.

In all but the two smallest rural districts, we had two field-work support staff.

Fieldworkers (sampling and data entry staff) were hired as described below.

#### *Fieldworker training and supervision*

We worked with consultants from the national statistics bureau to recruit fieldworkers from their network of partners in each sampling location. All had experience of collecting quantitative data using structured tools, for example in the national census or annual socio-economic survey rounds. All candidates attended an online training session to familiarise themselves with the aims and procedures of the study. One day prior to the start of fieldwork in each district, candidates attended a full-day face-to-face training covering general information about medicines and medicine quality; the purposes of the study and study procedures; data collection tools and equipment; study ethics; and financial and administrative issues. They were equipped with information about the conditions, symptoms, and side effects related to the medicines they were buying, and with scenarios to use when shopping. Candidates participated in simulations of all study procedures, including mystery shopping, sample processing and data entry (completing both the field and office data entry forms). Following the training, candidates were assigned specific roles in the study, either as a mystery shopper (also referred to as sample collectors), or as a data processor (data entry staff), in accordance with the aptitude they demonstrated during simulations. Sample collectors were given a daily advance for the purchase of medicines, and all fieldworkers received transport and communications costs, and were paid a daily wage.

Every morning during sampling, all fieldworkers attended a Zoom meeting with study supervisors. This allowed for troubleshooting of issues that arose in the field, and discussion of any changes to sampling plans. A STARmeds staff member at the hub was assigned to provide full-time support to data collectors by telephone and through a dedicated WhatsApp group during the sampling process. The site supervisor provided real-time support and problem-solving for data entry staff assigned to the hub.

#### *Data collection tools*

We collected data related to the samples using the open-source KoboCollect application, which runs on smart-phones, tablets and computers and uses Open Data Kit (ODK-)based forms. We designed three forms, one for sample collectors in the field, a second for online shopping, and a third for additional data entry at the hub.

To minimise data entry errors, forms were pre-populated with picklists, including names of outlets selected during sample selection, and all registered brands of the study medicines. Barcodes were entered using a scan function, and the location of outlet was collected using a geo-positioning function. Time spent collecting data, which was used in studies of customer convenience and to estimate the cost of post-market surveillance activities, was recorded automatically by the software data, using time-stamps. In addition, sample collectors noted the time spent in various tasks on their daily sample frames, which returned to the study hubs with each day's samples.

Modified examples of the ODK forms underlying the data collection tools can be downloaded for adaptation and reuse from our fieldwork Toolkit at https://doi.org/10.7910/DVN/OBIDHJ.

Data collected are shown in Table A8.

**Table A8: Key data collected using Kobo-collect.**

| Data point | Field form | Hub form |
|---|---|---|
| Medicine | Yes | Yes |
| Dosage | Yes | Yes |
| Formulation | Yes | Yes |
| Brand | Yes | Yes |
| Manufacturer | Yes | Yes |
| Batch number | No | Yes |
| Expiry date | No | Yes |
| Manufactured date | No | Yes |
| Type of medicine packaging | No | Yes |
| Photos of medicine packaging | No | Yes |
| Photos of batch number | No | Yes |
| Photos of expiry date | No | Yes |
| Number of samples | Yes | Yes |
| Registration Number | No | Yes |
| City | Yes | No |
| Location | Yes | No |
| Type of outlet | Yes | No |
| Outlet name | Yes | No |
| GIS data | Yes | No |
| Selling Price | Yes | No |
| Maximum Retail Price (HET) | No | Yes |
| Prescription required | Yes | No |
| Temperature at outlet | Yes | No |
| Time spent in sample collection | Yes | No |
| Time spent in data entry | Yes | Yes |

#### *Sample collection procedures*

For sampling from retail outlets, sampling staff were assigned a target price point (cheap or expensive) for the duration of the sampling period, and asked to dress appropriately for that price point (smartly for collectors of expensive medicines, and more simply for cheaper medicines). They were given personalised daily sample frames listing the selected outlets, the target medicines for the day, and a wallet card providing the target and minimum acceptable number of units per sample, as well as some common brand names of the medicine. Table A9 shows the number of units needed if all of the scheduled tests for the medicine ran to their final iteration in accordance with USP protocols, the ideal volume (included on prescriptions and requested by buyers), and the minimum acceptable volume (which allows for assay testing in duplicate, and a single round of dissolution).

**Table A9: Target, minimum number of units per medicine.**

| Medicine | Units needed for all tests* | Ideal units | Minimum accepted | Tests performed | | |
|---|---|---|---|---|---|---|
| | | | | Assay | Dissolution | Uniformity of content |
| Amlodipine | 64 tablets | 80 | 20 | Yes | Yes | Yes |
| Allopurinol | 44 tablets | 70 | 20 | Yes | Yes | No |
| Amoxicillin tablets | 34 tablets | 40 | 20 | Yes | Yes | No |
| Amoxicillin capsules | 44 tablets | 50 | 20 | Yes | Yes | No |
| Amoxicillin dry syrup | 5 60ml bottles | 5 | 3 | Yes | No | No |
| Cefixime | 44 capsules | 70 | 20 | Yes | Yes | No |
| Dexamethasone | 74 tablets | 80 | 20 | Yes | Yes | Yes |

*If all tests are repeated to the maximum iteration.

We aspired to ensure that all units in a sample were from the same batch number. However, since patients do not typically scrutinise batch numbers while buying medicines in a pharmacy, this was only feasible in the context of overt sampling.

Shoppers were provided with prescriptions for the target medicines, but instructed not to offer them unless requested by the sales staff. They were required to request a receipt.

Shoppers entered the shop and requested the medicines, using pre-prepared scenarios as necessary. These included buying medicines for a sick relative, or stocking up for a journey. They signalled desired price points using phrases such as "Is this the very best brand you have?" or "Do you have anything more affordable?"

On exiting the shop, they found a quiet place, and processed the sample as follows: Each sample (all strips or blisters of each unique medicine) was put in a separate Ziploc bag pre-labelled with a barcode. For each sample, shoppers opened a new KoboCollect form, scanned the barcode on the bag, recorded the location using geolocation function, entered medicine and price details, and uploaded the form to the server. Each sample Ziploc, which included additional barcode stickers for later processing, was stored in a daily sample bag labelled with the date and the fieldworker's name and study number. Daily sample bags were delivered to the hub in person or by courier at the end of the day.

If a target location did not have any brands of the target medicine (or had closed down or moved), shoppers noted this on data collection forms and called the hub for an alternative, randomly chosen outlet.

Staff collecting samples overtly from hospitals, health centres and health care providers followed the same data entry and sample handling procedures as mystery shoppers.

#### Online samples

Online shoppers (most of whom were STARmeds staff and colleagues from our academic group) were provided with sample frames including target online outlets, medicines and price points. They were requested to chat online with sellers to ascertain availability of brands and to verify prices, and to record orders made and price paid and delivery charges on a special data entry form. Samples were sent to a number of different addresses, and subsequently delivered to the data hub. STARmeds staff then assigned them a barcode, which was retrospectively matched with the entry on the online data-entry form. Online samples were then processed in the same way as other samples.

#### *Sample processing and storage*

##### Sample intake and packaging inspection

At the data hubs, each sample was entered on to a log sheet by barcode, and visually inspected by the site supervisor using a magnifying glass as necessary.

The site supervisor checked the primary packaging for integrity, as well as for any signs that may raise suspicions of falsification (such as misspellings, blurry printing, inconsistencies in typeface or direction of printing). Any suspicions were flagged on the log sheet for entry into the database. We did not have reference packaging from manufacturers, but over time built up a database of high-resolution photographs which in some cases allowed us to check and flag variations between products.

Where only part of a sample (only some strips, blisters or bottles) appeared suspicious and the supervisor judged that separate testing was warranted, the sample was split, and the suspect part assigned another barcode. More information on the specific signs and signals considered suspicious are provided in the study archive. We informed the national regulator BPOM in writing about any samples we judged to be highly likely to be falsified, proving batch numbers and other details. If we sampled these from active health care settings (doctor, midwife or health facility) we also immediately informed the health care provider.

Site supervisors also checked for uniformity of batches within a sample, and flagged samples with multiple batches to ensure that data processors entered the additional data.

##### Additional data entry

Data processors labelled each strip, blister or bottle in a sample with the additional barcode stickers included in the Ziploc. After scanning the barcode using dedicated tablets, they entered detailed data about the sample, including batch numbers, expiry dates and maximum retail prices. From within the KoboCollect application, they photographed the front of the primary packaging, and took close-ups of the batch number and expiry date.

##### Verification and storage

At the end of each day, another STARmeds staffer re-checked each sample against the log sheet (verifying medicine, dose, brand and batch numbers, and ensuring that all strips/blisters/bottles were correctly barcoded). Samples were separated by medicine and dose, ordered by barcode number, protected with bubble-wrap as necessary and stored in an airconditioned room in sealed plastic containers equipped with temperature loggers until the end of sampling, when they were hand carried or shipped by air cargo to Jakarta for testing. They were further stored in an airconditioned room in Jakarta until delivery to the laboratory.

#### *Sample triage*

When split samples were included, the absolute number of samples exceeded our projected testing budget. We therefore systematically excluded some samples from laboratory testing. These included samples where sample collectors aiming for different price points had bought the same product from the same outlet; samples where we had multiple examples of the same batch number of the same product from the same type of outlet in the same location; and samples which did not meet the minimum threshold for testing.

We pre-determined the order of laboratory tests for specific samples, according to the number of pills in each sample and the tests to be performed by medicine and formulation, as reported in the laboratory testing section.

Table A10 shows the number of samples excluded, the reasons for exclusion, and the total number sent for testing. The table also provides information on the actual numbers tested in each category; these differed from planned tests largely because products that failed assay were not processed for further tests, though in some cases also because of insufficiency of tablets.

**Table A10: Triage of samples for laboratory testing, and actual numbers tested**

| Testing Action | Criteria | Planned | Executed |
|---|---|---|---|
| **Total samples excluded** | | | **59** |
| Exclude from lab testing | Same product and same pharmacy<br>(Exclude one with fewest pills, else random) | 15 | |
| | <= 20 pills, and product sampled elsewhere | 6 | |
| | >=3 identical batches from different outlets but same regulated source type in same district<br>(Exclude one with fewest pills, else random) | 3 | |
| | >=2 identical products from different outlets but same regulated source type in same district<br>(Exclude one with fewest pills, else random) | 35 | |
| **Total samples sent for testing** | | | **1276** |
| Visual inspection | All samples | 1276 | 1276 |
| Assay testing only | <= 10 pills, unique or suspect product* | 30 | 55 |
| | All dry syrup formulations of amoxicillin** | 77 | 77 |
| Assay, dissolution and uniformity of content | Samples of amlodipine and dexamethasone with adequate pills to perform all three tests (to dissolution stage 3): at least one sample per product per district if available. Online samples not included. | 177 | 172 |
| Assay and dissolution only | All non-excluded solid form samples of allopurinol, amoxicillin and cefixime | 700 | 676 |
| | All remaining samples of amlodipine and dexamethasone | 293 | 292 |
| Assay and uniformity only | | 0 | 4 |

*Most of these are single suspect strips split from larger samples, e.g. because printing on this strip differs from the standard.
** This included two dry formulations of amoxicillin intended for injection, which were excluded from subsequent analysis

#### *Laboratory testing*

The only laboratory in Indonesia pre-qualified by WHO for the testing of medicine quality is BPOM's central testing laboratory, which was unable to process our samples because of COVID-19 related workload. Samples were tested at PT Equilab International, a private laboratory in Jakarta which is ISO/IEC 17025: 2017 certified (and is also WHO prequalified for bioavailability testing). STARmeds staff reviewed raw data as testing progressed, and worked with laboratory staff to troubleshoot any issues (for example data reporting formats) as they arose.

Samples were sent to the laboratory in three batches. They were tested between April 2022 and February 2023. Two samples contained at least some pills which were expired when sampled; a further 40 contained at least one batch which was expired at the time of the sample's final test date for any test. meaning that 3.7% of tested samples were expired at the time of the final test. 36 of these (2.8 of all samples tested) contained expired pills at the time of assay testing.

Laboratory staff downloaded temperature data from the data logger, then logged incoming samples by barcode, noting pills per sample and expiry dates, and the designated testing order.

Tests undertaken by medicine and formulation are shown in Table A9, and limits of acceptability are shown in table A13.

The workflow in the laboratory was as shown in Figure A2 (for allopurinol, amoxicillin tablets and capsules and cefixime) and Figure A3 (for amlodipine and dexamethasone). Amoxicillin dry syrup was visually inspected and tested for identification and quantity of assay only. To reduce the risk of bias in reading test results, samples were tested by staff other than those who undertook visual inspection and sample preparation.

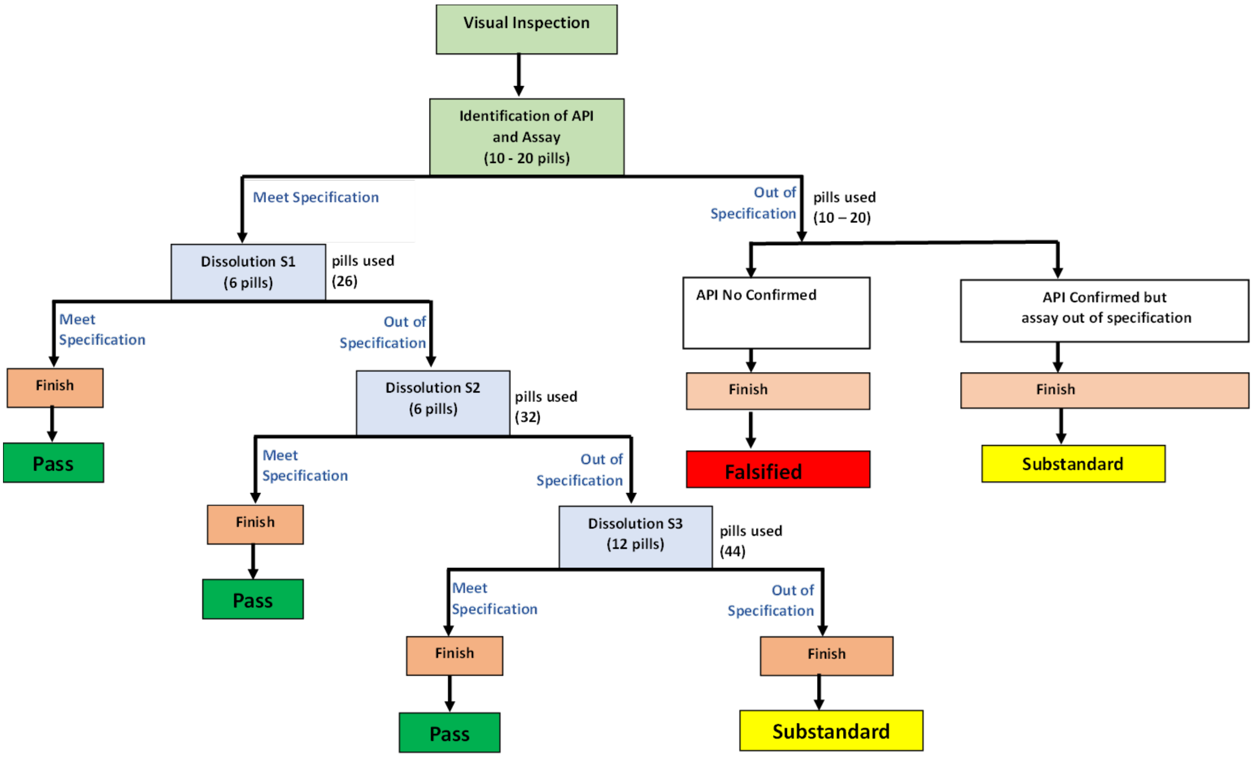


**Figure A2: Laboratory testing flow chart for sample at dosage ≥ 25 mg**

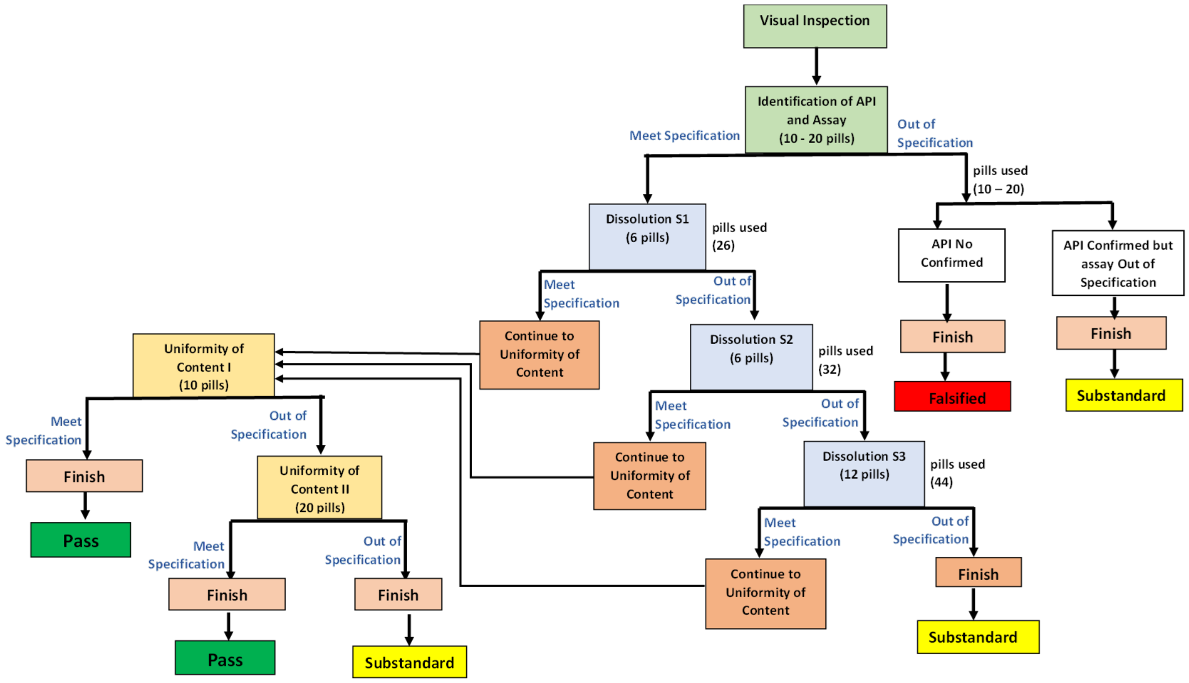


**Figure A3: Laboratory testing flow chart for sample at dosage < 25 mg**

All samples were tested using newly-imported United States Pharmacopeia (USP) reference standards, with valid certificates of analysis. All except for cefixime capsules were tested according to USP 43 NF38 monographs.(United States Pharmacopeia 2021) No USP monograph was available for cefixime capsules, so we used the Supplement 1 of *Farmakope Indonesia* 6$^{th}$ edition that refers to the Chinese Pharmacopeia methods.(Kementerian Kesehatan Republik Indonesia 2020)

Despite using USP/FI (and thus validated) methods, Equilab verified all methods before testing study samples, conducting within and between run repeatability testing for precision, accuracy and robustness. Full protocols for testing of all sample types are provided in the study archive. We did not conduct any inter-laboratory validation.

#### Appearance

After sample units were removed from primary packaging but before they were processed for identification testing, laboratory staff inspected them visually, noting colour, markings and any unexpected features such as cracking or crumbling.

#### Sample preparation

Sample preparation for each medicine and formulation is described in the detailed laboratory protocols provided in the study archive. For allopurinol, cefixime, dexamethasone and capsule formulations of amoxicillin, 20 tablets were powdered in order to prepare the sample solution. For amlodipine and amoxicillin, 5 tablets were dissolved directly in solvent. The 2.5 litre flasks necessary for the one-step dissolution of 500mg amoxicillin tablets are not available in Indonesia. We initially used a two-step process in which 5 tablets were dissolved in one litre of solvent; 2mls of this solution were drawn off, then further diluted to achieve the necessary concentration of 1mg/ml. Early monitoring indicated this impeded correct dissolution. We then adopted the following approach:

- Dissolve two tablets in one litre of buffer
- Dissolve a further two tablets in one litre of buffer
- Dissolve one tablet in 500ml of buffer
- Mix the three solutions; take a portion for centrifuge and filtering
- Draw off aliquot of 1ml for testing.

For consistency, all samples were retested using this approach, which is the basis of reported results.

#### Identification

The presence of the expected active ingredient was demonstrated by comparing the dominant peaks in the chromatograms of the sample solution and that of the reference standard. If the retention time for the dominant peaks corresponded, identification was considered positive.

#### Assay testing

Assay testing was conducted following the procedures in each monograph.

For all molecules, assay testing was done by high-performance liquid chromatography (HPLC -UV Waters, Aliance 2695 with UV Detector 2489), using chromatographic conditions as specified by USP.

Assay testing was conducted in duplicate for each sample. A sample was considered out of specification if the average of the two tests fell outside of the limits given in Table A13.

##### Uniformity of content (also known as uniformity of dosage units)

Uniformity of content was conducted for amlodipine and dexamethasone (Table A9). Each of 10 tablets was weighed and dissolved individually. Each was sonicated and stirred to disintegrate. Dexamethasone was filtered for injection into the HPLC machine, while for amlodipine absorbance was measured by spectrophotometer. The % of labelled active ingredient was measured separately for each unit. The acceptance value was calculated according to the formula given by USP, in its General Chapter 905 on uniformity of dosage units.(United States Pharmacopeia 2021) This formula takes into account individual assay values and the standard deviation between them. The uniformity of content was considered in specification if the resulting acceptance value was $\leq$ 15.0 and no individual tablet fell outside of the limits set by the formula.

If the acceptance value exceeded 15.0 at the first round of uniformity testing, the assay value of a further 20 tablets were measured individually. The USP formula was used to calculate the acceptance value across all 30 tablets measured. The uniformity of content was considered in specification if the resulting acceptance value was $\leq$ 15.0 and no individual tablet fell outside of the limits set by the formula.

#### Dissolution testing

Dissolution testing was conducted following the procedures in each monograph, using the dissolution apparatus recommended by USP, set to the speed -- revolutions per minute or rpm -- required for the specific active ingredient, as shown in Table A11. Tablets and capsules were dissolved at a temperature of 37 ± 0.5 ºC.

Dissolution of allopurinol, amlodipine, amoxicillin capsules and cefixime was analysed using Spectrophotometer-UV/VIS (Shimadzu UV-1800). Amoxicillin tablets and dexamethasone were analysed using HPLC. No dissolution testing was performed on amoxicillin dry syrup formulation.

**Table A11: Equipment and parameters for dissolution testing**

| Medicine | Apparatus, and RPM | Medium; volume | Time* | Analytical method |
|---|---|---|---|---|
| Amlodipine | USP 2 Paddle, 75 rpm | 0.01 N HCl; 500 mL | 30 | Spectrophotometric |
| Dexamethasone | USP 1 Basket, 100 rpm | 0.1 N HCl; 500 mL | 30 | Chromatographic** |
| Allopurinol | USP 2 Paddle, 75 rpm | 0.01 N HCl; 900 mL | 45 | Spectrophotometer |
| Amoxicillin tablet | USP 2 Paddle, 75 rpm | Water; 900 mL | 30 | Chromatographic |
| Amoxicillin capsule | USP 1 Basket, 100 rpm (for 250 mg capsules<br>USP 2 Paddle, 75 rpm (for 500 mg capsules) | Water; 900 mL | 60 | Spectrophotometric |
| Cefixime tablet | USP 1 Basket, 100 rpm | Phosphate buffer pH 7.2, 900 mL | 45 | Spectrophotometric |
| Cefixime capsule | USP 1 Basket, 100 rpm | Phosphate buffer pH 7.2, 900 mL | 45 | Spectrophotometric |

*In minutes **HPLC

Dissolution was conducted in 3 stages (S1, S2, S3) as necessary. At S1 and S2, 6 tablets were used. At S3, 12 tablets were used. Table A12 shows the acceptance criteria for dissolution, and our workflow.

**Table A12: Workflow for dissolution testing, and acceptance criteria**

| Stage | Units | Criteria | Result | Action & final result |
|---|---|---|---|---|
| 1 | 6 | Any unit < Q-25? | Yes | Finish; Failed |
| | | Any 2 units <Q-15 | Yes | Finish; Failed |
| | | Any unit <Q+5 | No | Finish; Passed |
| | | | Yes | Continue to Stage 2 |
| 2 | 6 | Any unit < Q-25? | Yes | Finish; Failed |
| | | Any 2 of 12 units (S1&S2) <Q-15? | Yes | Finish; Failed |
| | | Average of 12 units (S1&S2) <Q | No | Finish; Passed |
| | | | Yes | Continue to Stage 3 |
| 3 | 12 | Any unit < Q-25? | Yes | Finish; Failed |
| | | Any 2 of 24 units (S1&S2&S3) <Q-15? | Yes | Finish; Failed |
| | | Average of 24 units (S1&S2&S3) <Q | No | Finish; Passed |
| | | | Yes | Finish; Failed |

#### Impurity testing

Our budget did not stretch to testing for impurities for any study samples.

##### Microbiological testing

We performed microbiological testing on all samples of antibiotics for which we had sufficient tablets/capsules/bottles remaining after other tests were concluded (n=321).

Both amoxicillin and cefixime were tested against *Escherichia coli* ATCC 25922, and amoxicillin was additionally tested against *Staphylococcus aureus* ATCC 25923, using the Kirby-Bauer disc diffusion method as described in the Clinical Laboratory Standard Institute's 30th Edition.(Clinical & Laboratory Standers Institute 2020)

#### Verification of authenticity

We sought to verify with market authorisation holders that the combination of batch numbers and expiry dates of the samples we collected matched with their production records. After receiving laboratory results, we wrote to all market authorisation holders of any product tested in our study (n= 79) both electronically and in hard copy. We provided them with the batch numbers and expiry dates of all of their samples, as well as high-resolution photographs of the primary and (if available) secondary packaging, and asked them to check production records and packaging for any anomalies.

If the market authorisation holder confirmed a product was falsified (for example, because the expiry date did not match the batch number, possibly indicating extension of expiry dates by falsifiers), we categorised it as 'confirmed falsified'.

#### Quality definitions

In identifying the active ingredient, Table A13 shows the limits within which measures for study medicines must fall to meet quality specification in both USP and Farmakope Indonesia (for cefixime capsules, Supplement 1 of Farmakope Indonesia 6$^{th}$ ed referring to the Chinese Pharmacopeia method).

In microbiological testing, we used the breakpoints defined by Clinical Laboratory Standards Institute (CLSI) 30th edition.

**Table A13: USP Acceptability limits for medicines in the study, USP and Farmakope Indonesia**

| Active ingredient | Identification | Assay (%) | Dissolution (%) ('Q') | Content Uniformity | Microbiology** | |
|---|---|---|---|---|---|---|
| | | | | | *E. Coli* | *S. Aureus* |
| Allopurinol | The retention time of the major peak of the sample solution corresponds to that of the reference solution | 93.0 – 107.0 | 75% in 45 minutes | NA | NA | NA |
| Amoxicillin, tablet | | 90.0 - 120.0 | 75% in 30 minutes | NA | Inhibitory zone >=14 mm | Inhibitory zone >=26 mm |
| Amoxicillin, capsule | | 90.0 - 120.0 | 80% in 60 minutes | NA | | |
| Amoxicillin, dry syrup | | 90.0 - 120.0 | NA | NA | | |
| Cefixime, tablet | | 90.0 – 110.0 | 75% in 45 minutes | NA | Inhibitory zone >=17 mm | NA |
| Cefixime, capsule* | | 90.0 – 110.0 | 80% in 45 minutes | NA | | NA |
| Amlodipine | | 90.0 – 110.0 | 75% in 30 minutes | Acceptance value ⩽ 15.0, and no individual tablet has an assay value that falls outside USP-specified limits. | NA | NA |
| Dexamethasone | | 90.0 - 110.0 | 80% in 30 minutes | | NA | NA |

NA: Not applicable

*No USP monograph available

** Clinical and Laboratory Standards Institute antimicrobial susceptibility testing standards

We considered a sample out of specification if it failed at least one of the chemical tests to which it was subjected. This definition is used throughout our papers, unless they state otherwise. Samples that did not have sufficient tablets to conduct all rounds of testing were not considered out of specification (n=17), unless they also failed on a different parameter (n=1).

Our approach reflects the likelihood that in resource-constrained settings, patients care more about whether a medicine "works" -- delivers the expected amount of active ingredient, as reflected in assay and dissolution tests -- than about technicalities related to registration status, expiry date or packaging. Researchers wishing to use a different definition are free to recalculate from the sample-level data, which include all raw laboratory outcomes and are posted in the study archive.

##### Substandard versus falsified medicines

We attempted to distinguish between substandard and falsified products by reporting pharmacopeial test results to all manufacturers, giving them the opportunity to verify whether products were genuine. We classified the results as shown in Table A14

**Table A14: Classification of substandard and falsified medicines as reported in results**

| | **Failed any test** | **Passed all tests** |
|---|---|---|
| Conforms with manufacturing records | Probable substandard | Quality product |
| Does not conform with manufacturing records | Falsified | Falsified |
| No response from manufacturer | Probable substandard | Quality product |

### Data management

Data from the online and offline sample purchasing forms were merged on barcode with data collected using the additional field data entry form, and checked for completeness and consistency. Missing data or discrepancies were resolved by study staff in the field with reference to processed samples and data collectors' daily forms, or later in the main study office with reference to package photographs and purchase receipts. All data cleaning was performed using Stata 17, for record-keeping and reproducibility. Data analysis was performed using Stata 18.

Cleaned sample data were merged with laboratory data on barcode.

### Ethics and reporting

#### *Ethical and policy review*

During the design phase, the study was discussed extensively with the Indonesian medicine regulator BPOM, a multisectoral national working group on medicine quality, and an external academic advisory board. The study protocol was approved by institutional review boards at Universitas Indonesia and Imperial College London.

The reference number for permits from Universitas Indonesia was 970/UN2.F1/ETIK/PPM.00.02/2020, 7 September 2020, with an extension letter on September 13th, 2021; S-736/UN2.F1/ETIK/PPM.00.02/2021. Imperial College Research Ethics Committee approved the study with reference number 21IC7265 on 16 November 2021. We also explained the aims and methods of the study to the district governments of the eight districts in which samples were collected, and obtained a letter of approval from each.

While sampling, we identified some samples that we suspected of falsification. We notified BPOM (and in cases of strongest suspicion manufacturers) while the study was still ongoing, providing details and high resolution photographs. Where suspect samples came from health care providers dispensing directly to patients, we also notified them immediately, allowing them to quarantine potentially dangerous products while awaiting regulatory follow-up.

When testing was completed, we provided details to all manufacturers, asking them to confirm that batch number/expiry date combinations were consistent with their records, as described above. After the deadline for response passed, we provided sample level raw laboratory data to those who responded, for both suspect and non-suspect samples.

##### Health protocols

Samples were collected during the COVID-19 pandemic. We required (and paid for) PCR tests for everybody planning to attend the face-to-face trainings. Those with positive tests were excluded. All STARmeds staff and fieldworkers followed full health protocols, including wearing masks, during sampling, sample processing, and data entry.

##### MEDQUARG

We have reported the study methods and results as closely as possible according to the MEDQUARG guidelines (see Table 1 of 10.1371/journal.pmed.1000052). Our reporting checklist is provided in the study archive.

#### Reporting raw prevalence of poor quality medicines

Raw estimates of prevalence of out of specification medicines were calculated by dividing the number of samples failing at least one chemical test by the total number of samples tested, using data from the STARmeds study alone [4]. We excluded microbiological test results from our quality definitions because we were only able to perform them on the sub-set of samples with sufficient remaining pills, creating a possibility of bias.

In order to examine the effect of including additional quality parameters, we also calculated prevalence by two additional measures, which we refer to as Expanded, and Maximum. The parameters included in the denominator of each of the three measures are summarised in Table A15.

**Table A15: Definitions of quality use in analysis**

| | **Measure of quality** | **Core** | **Expanded** | **Maximum** |
|---|---|---|---|---|
| | Failed assay testing | **Yes** | **Yes** | **Yes** |
| OR | Failed dissolution testing | **Yes** | **Yes** | **Yes** |
| OR | Failed uniformity of content testing | **Yes** | **Yes** | **Yes** |
| OR | Expired at time of purchase, | **No** | **Yes** | **Yes** |
| OR | No expiry date on primary packaging | **No** | **Yes** | **Yes** |

| | | | | |
|---|---|---|---|---|
| OR | Broken or damaged at time of purchase | No | Yes | Yes |
| OR | Unlicensed in the local market | No | Yes | Yes |
| OR | Confirmed falsified by market authorisation holder | No | No | Yes |

We compared raw prevalence across medicines, geographies and sources of samples, testing for statistically significant differences using Pearson's Chi squared test, deeming differences significant at the 95% level.

### Adjusted estimates of prevalence of poor quality medicines

A major aim of our study was to compare raw estimates of substandard medicines with estimates adjusted for the likelihood that a patient would actually take that specific product and brand. Here we describe the methods for weighting data by market size to yield adjusted estimates.

#### *Data sources*

The universe of products for the study medicines was defined by the BPOM registration database (Table A1: [1]). Test results came from the STARmeds study (Table A1: [4]), while volume data were taken from IQVIA sales volume for calendar 2022 (Table A1: [3]), supplemented by data from the national medicine procurement system (Table A1: [2]). A total of 262 individual products (unique in terms of medicine, formulation, dosage and brand or -- for unbranded products -- market authorisation holder) were sampled by both IQVIA, in their routine market surveys, and STARmeds.

If a product was listed by BPOM but registered no sales on the public procurement platform and was not found in the market by either IQVIA or STARmeds, we assumed that it was a dormant registration, and dropped it from the estimates. For registered products that were sampled in STARmeds but not listed by IQVIA, we imputed volumes systematically as described below. All assumptions used in a draft version of the imputation model were discussed with the national technical working group on medicine quality estimation, which includes representatives from the medicine regulator, other health agencies, industry, professional associations and academia. The methods below incorporate their consensus on the most reasonable methods and assumptions for imputation.

##### Product volume

IQVIA data on market volumes in Indonesia are derived from a panel of 250 hospitals, 1064 pharmacies and 250 non-prescription medicine shops, sampled to be nationally representative, and audited quarterly. Volumes are reported separately by quarter and type of outlet. The hospitals include those in the public and the private sectors. Other public health facilities, including both public and private primary care centres, are NOT captured in IQVIA data.

IQVIA listings included a few products (market authorisation holders, brands, active ingredients and dosages) which are not authorised in the Indonesian market. These may result from legacy data or other errors; volumes of these products were generally small and they were dropped from the estimations. The data also included a significant volume of products classified as "generic manufacturer" -- 31% by volume over the five study medicines.

Individual brand and market authorisation data are not available for these products. IQVIA clarified that these represented products which were listed only by active ingredient name in hospital or pharmacy inventory systems, and were likely to come from market authorisation holders already captured in the record-keeping systems of other outlets with more detailed data.

There were also products sampled in STARmeds that were not listed in IQVIA data (n=18). We assume these products are relatively rare in the Indonesian market. We imputed volumes for these products as described in Step 4 below.

We estimated market volume by product as follows:

**Step 1**: We combined data across four quarters to estimate IQVIA-recorded volumes for calendar 2022 by product (active ingredient, brand, dose and formulation).

**Step 2**: For each product, we combined data for the pharmacy and medicine shop channels to represent the retail sector. These data should also include all online sales volumes from regulated and semi-regulated platforms/sources, since these are sourced from registered physical pharmacies.

**Step 3**: For the study products that were winners of public procurement tenders in 2021 (and thus procured and distributed in 2022; n=17), we added realised sales volumes from the public procurement platforms to the retail sales volumes recorded by IQVIA. This will have captured the bulk of medicines provided in primary health centres. Most hospitals buying these products would also have sourced them through the public procurement platform. For these products, we excluded volumes in IQVIA's hospital audit data, to avoid double counting units already captured in the public procurement volumes.

**Step 4**: For products (active ingredient, brand, formulation, and dose) that have a valid market authorisation with BPOM and were sampled by STARmeds but which were not listed by IQVIA (n=18), we imputed missing volumes as follows:

- If volumes are available for a different dose of the same active ingredient, formulation and brand: Calculate ratio of volumes by dose for this medicine across the Indonesian market (e.g., ratio of amlodipine 5mg to amlodipine 10mg). Multiply volume of existing dose by appropriate ratio to estimate volume for missing dose.
- If no volume is available for the brand: use the median volume for all the registered study products which are listed in IQVIA but which were not sampled by STARmeds (n=113). The rationale for this is that we assume the products that were missing from either STARmeds or IQVIA sampling are both relatively rare in the Indonesian market. Volume data for those in IQVIA but not in STARmeds may thus be a good proxy for those found by STARmeds but not sampled by IQVIA.

**Step 5**: We distributed the significant volume of products listed by IQVIA as registered to "generic manuf." in proportion to the relative market size for each of the individual unbranded generic products in the Indonesian market.

**Step 6**: We did not adjust for volumes of online sales of either regulated online stores or regulated pharmacies selling online, since these should be captured in IQVIA data based on retail pharmacy stock records. We did, however, estimate sales through unregulated internet sellers by looking at their page rankings and sales figures on the general internet marketplaces where they operate. In our mapping searches, we learned three things relevant to these estimates:

- For medicines not subject to off-label use, unregulated sellers offer more branded than unbranded medicines;
- They usually appear very far down the page rankings when ordered by the default "closest match" and also "cheapest", implying that buyers have to scroll through hundreds of offers from verified pharmacies before finding an offer from an unregulated seller;
- Their platform-registered sales volumes are usually in single figures (and often zero), compared with the thousands or hundreds registered by most sellers on the first pages (by default page order).

This implies that customers have to work hard to buy from unverified sellers, something that we confirmed during our own sampling. For simplicity, we estimated additional volumes sold through unregulated internet sources as 0.01% of the retail sales volume for branded products, and 0.001% for unbranded products.

#### Sample weights

Our study randomised at the level of the outlet, but this imperfectly reflects the national market for study medicines. We could not randomised at the level of the individual medicine, because it is not feasible to get a complete listing of volumes of every study medicine at every outlet using a mystery shopped method. Private hospitals were selected randomly, but, at just one per district, were comparatively under-sampled relative to retail outlets in all but the smallest districts. In addition, in our internet sampling we deliberately sought out hard-to-find "wild type" sellers (individuals on general internet marketplaces or social media or other unauthorised sellers not linked with a regulated pharmacy), and targeted a variety of brands and products.

To allow for comparisons between medicines, we further aimed to test the same number of samples of each target medicine and dose, despite large differences in the overall volumes of the medicines in the Indonesian market.

To correct for these differences, we weighted our data to reflect a product's weight in the market, relative to its weight in our sample. We calculated weights as follows:

**Step 1: Calculate the *Universe of 5 medicines*** *[variable name: total_volume]*
Sum all of the reported or imputed volumes for all five medicines included in the study, including non-targeted doses, but excluding formulations such as injectable versions not included in our sampling.

**Step 2: Calculate *Market shares***
For each individual product in the market, calculate market share in the physical, regulated and semi-regulated online market ["Non-wild" market share], and for unregulated internet sales by individuals ["Wild-type" market share] as follows:

Volume of product in physical and linked online outlets/Universe of 5 medicines
*[variable name: market_share_nw]*

AND

Volume of product sold by individuals on unregulated platforms/Universe of 5 medicines.
*[variable name: market_share_w]*

**Step 3: Calculate *Study shares*** *[variable name: prop_tested]*
For each individual product in the study sample, calculate the proportion of samples collected in the physical, regulated and semi-regulated online market [Study share, non-wild samples],

and those bought from unregulated internet sales by individuals [Study share, wild-type samples] as follows:

Number of samples of this product from regular source/Total number of samples

AND

Number of samples of this product from wild-type source/Total number of samples.

Step 4: **Calculate the *Adjustment weight*** *[variable name: weight]*

For each sample in the study, calculate a weight as follows:

Market share/Study share

using the Regular or Wild-type values as appropriate to the source of the specific sample.

##### Within-medicine weights

For estimates of prevalence of poor quality medicines within each medicine (rather than across the universe of 5 study medicines), we re-calculated the universe, market share, study share and adjustment weights per active ingredient. *[Variable names as above, with the extension "_mol"]*

##### Weighted analysis

Data were re-analysed using these adjustment weights, which correct for over- or under-sampling relative to national market share. We used Stata 18's survey estimation commands, setting our weight variable [*weight]* as the pweight. The adjusted sample size was 1192 (compared with an unweighted sample size of 1274), reflecting the oversampling of a number of brands relative to their small share of the market.

#### Availability of data and code

Study data, including granular laboratory results, are available in the STARmeds repository, at https://dataverse.harvard.edu/dataverse/STARmeds. Company, brand and granular source data are masked in accordance with the requirements of our ethical approval. The masking codes are consistent between variables (so for example the brand "allo a00286_inn" is an unbranded allopurinol product marketed by market authorisation holder "a00286"). These data may be freely downloaded and used by other researchers under a CC BY-NC-SA 4.0 license that allows for non-commercial use and requires users to make any incorporating/ resulting work available on similar terms.

The Stata-format code used to estimate and impute volumes and weight the data are also available in the repository.

#### Study Group members, and acknowledgements

STARmeds was a collaboration between Universitas Pancasila, Imperial College London and Erasmus University Rotterdam.

STARmeds study group members listed alphabetically by institution. Group member roles are provided in the supplementary materials for each individual papers. Joint Principal Investigators are shown in bold.

**Universitas Pancasila**

**Yusi Anggriani**, Esti Mulatsari, William Nathanial, Yunita Nugrahani, **Elizabeth Pisani**, Jenny Pontoan, Ayu Rahmawati, Mawaddati Rahmi, Stanley Saputra, Hesty Utami.

**Imperial College London**

Adrian Gheorghe, **Katharina Hauck**, Sarah Njenga, Sara Valente de Almeida.

**Erasmus University Rotterdam**

Amalia Hasnida.

The team would like to thank Indonesia's medicine regulator Badan Pengawas Obat dan Makanan and Statistics Indonesia for active support in developing the methods described here. The majority of the data collectors were partners of the national statistics bureau (Badan Pusat Statistik). We thank them for their hard work. We thank BPS statistical consultants (Ardi Adji and Budi Santoso).

We also thank members of a multisectoral working group on medicine quality estimation known as PEMO, which groups 12 Indonesian government institutions and 5 professional or industry associations, for advice provided over the course of the study, as well as the members of our Study Advisory Group (Michael Deats, Kharisma Nugroho, Yodi Mahendradhata, Raffaella Ravinetto, Selma Siahaan, Val Snewin, Virginia Wiseman and Firman Witoelar) for useful advice.

United States Pharmacopeia provided reference standards at discounted prices. The study was funded by UK taxpayers through the UK Department of Health and Social Care and the National Institute for Health Research. We are grateful to them all.

United States Pharmacopeia. 2021. “United States Pharmacopeia National Formulary (43rd Edition NF38).” USP.

WHO Expert Committee on Specifications for Pharmaceutical Preparations. 2016. “Guidelines on the Conduct of Surveys of the Quality of Medicines. WHO Technical Report Series, No. 996, 2016, Annex 7.” World Health Organization. https://digicollections.net/medicinedocs/#d/s22404en.

World Health Organization. 2017. “A Study on the Public Health and Socioeconomic Impact of Substandard and Falsified Medical Products.” WHO/EMP/RHT/2017.02. Geneva: WHO. http://who.int/medicines/regulation/ssffc/publications/Layout-SEstudy-WEB.pdf.
