## Supplementary Table 1 for "A randomised survey of the quality of antibiotics and other essential medicines in Indonesia, with volume-adjusted estimates of the prevalence of substandard medicines"

| Medicine | Dose and forumulation | MA holder | Brand | Samples from physical sour online sales by verified p | |
|---|---|---|---|---|---|
| | | | | Market share | Share of STARmeds samples |
| allopurinol | 100_tab | a00309 | allo a00309_inn | 4.76 | 2.59 |
| allopurinol | 100_tab | a00215 | allo a1407 | 0.63 | 1.49 |
| allopurinol | 100_tab | a00286 | allo a00286_inn | 0.29 | 0.31 |
| allopurinol | 100_tab | a00196 | allo a3149 | 0.27 | 1.65 |
| allopurinol | 100_tab | a00003 | allo a00003_inn | 0.27 | 0.63 |
| allopurinol | 100_tab | a00385 | allo a00385_inn | 0.16 | 1.81 |
| allopurinol | 100_tab | a00232 | allo a00232_inn | 0.15 | 0.31 |
| allopurinol | 100_tab | a00246 | allo a00246_inn | 0.11 | 0.55 |
| allopurinol | 100_tab | a00082 | allo a3209 | 0.08 | 1.18 |
| allopurinol | 100_tab | a00333 | allo a0886 | 0.04 | 0.47 |
| allopurinol | 100_tab | a00004 | allo a2492 | 0.03 | 0.47 |
| allopurinol | 100_tab | a00438 | allo a4833 | 0.03 | 0.39 |
| allopurinol | 100_tab | a00309 | allo a4371 | 0.02 | 0.16 |
| allopurinol | 100_tab | a00061 | allo a00061_inn | 0.01 | 0.08 |
| allopurinol | 100_tab | a00068 | allo a5415 | 0.01 | 0.31 |
| allopurinol | 100_tab | a00215 | allo a00215_inn | 0.01 | 0.31 |
| allopurinol | 100_tab | a00334 | allo a00334_inn | 0.01 | 0.63 |
| allopurinol | 100_tab | a00382 | allo a00382_inn | 0.01 | 0.16 |
| allopurinol | 100_tab | a00295 | allo a00295_inn | 0.01 | 0.08 |
| allopurinol | 100_tab | a00130 | allo a00130_inn | 0.01 | 0.39 |
| allopurinol | 100_tab | a00270 | allo a0353 | 0.00 | 0.08 |
| allopurinol | 100_tab | a00204 | allo a1667 | 0.00 | 0.08 |
| allopurinol | 100_tab | a00076 | allo a3077 | 0.00 | 0.31 |
| allopurinol | 100_tab | a00286 | allo a2109 | 0.00 | 0.16 |
| allopurinol | 100_tab | a00144 | allo a1017 | 0.00 | 0.00 |
| allopurinol | 100_tab | a00367 | allo a00367_inn | 0.00 | 0.08 |
| allopurinol | 100_tab | a00387 | allo a0254 | 0.00 | 0.08 |
| allopurinol | 100_tab | a00096 | allo a00096_inn | 0.00 | 0.08 |
| allopurinol | 100_tab | a00085 | allo a2743 | 0.00 | 0.00 |
| allopurinol | 100_tab | a00405 | allo a00405_inn | 0.00 | 0.08 |
| allopurinol | 100_tab | a00130 | allo a5069 | 0.00 | 0.16 |
| allopurinol | 100_tab | a00234 | allo a00234_inn | 0.00 | 0.16 |
| allopurinol | 100_tab | a00405 | allo a3239 | 0.00 | 0.08 |
| allopurinol | 100_tab | a00110 | allo a5364 | 0.00 | 0.00 |
| allopurinol | 100_tab | a00094 | allo a1651 | 0.00 | 0.08 |
| allopurinol | 300_tab | a00309 | allo a00309_inn | 1.46 | 1.41 |
| allopurinol | 300_tab | a00286 | allo a00286_inn | 0.37 | 0.63 |
| allopurinol | 300_tab | a00215 | allo a1407 | 0.32 | 0.39 |
| allopurinol | 300_tab | a00196 | allo a3149 | 0.20 | 0.24 |
| allopurinol | 300_tab | a00082 | allo a3209 | 0.10 | 0.24 |
| allopurinol | 300_tab | a00102 | allo a5618 | 0.07 | 0.24 |
| allopurinol | 300_tab | a00004 | allo a2492 | 0.03 | 0.31 |
| allopurinol | 300_tab | a00003 | allo a00003_inn | 0.03 | 0.08 |
| allopurinol | 300_tab | a00232 | allo a00232_inn | 0.03 | 0.24 |
| allopurinol | 300_tab | a00333 | allo a0886 | 0.02 | 0.16 |
| allopurinol | 300_tab | a00061 | allo a0206 | 0.02 | 0.16 |

| | | | | | |
|---|---|---|---|---|---|
| allopurinol | 300_tab | a00309 | allo a4371 | 0.01 | 0.00 |
| allopurinol | 300_tab | a00286 | allo a2109 | 0.01 | 0.00 |
| allopurinol | 300_tab | a00244 | allo a2179 | 0.01 | 0.00 |
| allopurinol | 300_tab | a00270 | allo a0353 | 0.01 | 0.00 |
| allopurinol | 300_tab | a00438 | allo a4833 | 0.01 | 0.16 |
| allopurinol | 300_tab | a00385 | allo a00385_inn | 0.00 | 0.00 |
| allopurinol | 300_tab | a00347 | allo a2254 | 0.00 | 0.00 |
| allopurinol | 300_tab | a00204 | allo a1667 | 0.00 | 0.08 |
| allopurinol | 300_tab | a00144 | allo a1017 | 0.00 | 0.16 |
| allopurinol | 300_tab | a00439 | allo a1345 | 0.00 | 0.08 |
| allopurinol | 300_tab | a00110 | allo a5364 | 0.00 | 0.00 |
| amlodipine | 10_tab | a00061 | amlo a00061_inn | 5.89 | 0.24 |
| amlodipine | 10_tab | a00214 | amlo a00214_inn | 2.55 | 0.16 |
| amlodipine | 10_tab | a00286 | amlo a00286_inn | 1.87 | 0.08 |
| amlodipine | 10_tab | a00309 | amlo a00309_inn | 1.34 | 0.16 |
| amlodipine | 10_tab | a00311 | amlo a00311_inn | 1.10 | 0.08 |
| amlodipine | 10_tab | a00149 | amlo a00149_inn | 0.99 | 0.16 |
| amlodipine | 10_tab | a00010 | amlo a00010_inn | 0.64 | 0.08 |
| amlodipine | 10_tab | a00116 | amlo a00116_inn | 0.50 | 0.08 |
| amlodipine | 10_tab | a00285 | amlo a00285_inn | 0.10 | 0.08 |
| amlodipine | 10_tab | a00402 | amlo a1778 | 0.09 | 0.24 |
| amlodipine | 10_tab | a00334 | amlo a00334_inn | 0.07 | 0.08 |
| amlodipine | 10_tab | a00116 | amlo a2350 | 0.07 | 0.00 |
| amlodipine | 10_tab | a00130 | amlo a00130_inn | 0.06 | 0.08 |
| amlodipine | 10_tab | a00140 | amlo a3115 | 0.03 | 0.08 |
| amlodipine | 10_tab | a00328 | amlo a3739 | 0.03 | 0.00 |
| amlodipine | 10_tab | a00144 | amlo a00144_inn | 0.02 | 0.08 |
| amlodipine | 10_tab | a00102 | amlo a2363 | 0.02 | 0.00 |
| amlodipine | 10_tab | a00412 | amlo a1815 | 0.01 | 0.00 |
| amlodipine | 10_tab | a00199 | amlo a4380 | 0.00 | 0.00 |
| amlodipine | 10_tab | a00155 | amlo a0913 | 0.00 | 0.00 |
| amlodipine | 10_tab | a00141 | amlo a00141_inn | 0.00 | 0.08 |
| amlodipine | 5_tab | a00214 | amlo a00214_inn | 3.08 | 1.33 |
| amlodipine | 5_tab | a00149 | amlo a00149_inn | 2.86 | 1.41 |
| amlodipine | 5_tab | a00309 | amlo a00309_inn | 2.35 | 2.20 |
| amlodipine | 5_tab | a00311 | amlo a00311_inn | 1.60 | 0.39 |
| amlodipine | 5_tab | a00116 | amlo a00116_inn | 0.97 | 0.47 |
| amlodipine | 5_tab | a00286 | amlo a00286_inn | 0.71 | 0.39 |
| amlodipine | 5_tab | a00010 | amlo a00010_inn | 0.46 | 0.24 |
| amlodipine | 5_tab | a00402 | amlo a1778 | 0.37 | 0.94 |
| amlodipine | 5_tab | a00215 | amlo a4212 | 0.20 | 1.02 |
| amlodipine | 5_tab | a00116 | amlo a2350 | 0.17 | 0.47 |
| amlodipine | 5_tab | a00347 | amlo a00347_inn | 0.12 | 0.08 |
| amlodipine | 5_tab | a00021 | amlo a4657 | 0.08 | 0.24 |
| amlodipine | 5_tab | a00140 | amlo a3115 | 0.07 | 0.24 |
| amlodipine | 5_tab | a00246 | amlo a00246_inn | 0.07 | 0.08 |
| amlodipine | 5_tab | a00405 | amlo a00405_inn | 0.06 | 0.16 |
| amlodipine | 5_tab | a00285 | amlo a00285_inn | 0.05 | 0.08 |
| amlodipine | 5_tab | a00328 | amlo a3739 | 0.05 | 0.63 |
| amlodipine | 5_tab | a00334 | amlo a00334_inn | 0.05 | 0.63 |
| amlodipine | 5_tab | a00333 | amlo a4553 | 0.04 | 0.16 |
| amlodipine | 5_tab | a00121 | amlo a1686 | 0.04 | 0.08 |

| | | | | | |
|---|---|---|---|---|---|
| amlodipine | 5_tab | a00068 | amlo a4739 | 0.04 | 0.24 |
| amlodipine | 5_tab | a00102 | amlo a2363 | 0.03 | 0.31 |
| amlodipine | 5_tab | a00061 | amlo a00061_inn | 0.03 | 0.47 |
| amlodipine | 5_tab | a00412 | amlo a1815 | 0.03 | 0.08 |
| amlodipine | 5_tab | a00231 | amlo a4734 | 0.03 | 0.08 |
| amlodipine | 5_tab | a00294 | amlo a00294_inn | 0.02 | 0.16 |
| amlodipine | 5_tab | a00382 | amlo a00382_inn | 0.02 | 0.08 |
| amlodipine | 5_tab | a00381 | amlo a00381_inn | 0.02 | 0.08 |
| amlodipine | 5_tab | a00286 | amlo a4372 | 0.01 | 0.08 |
| amlodipine | 5_tab | a00119 | amlo a1270 | 0.01 | 0.00 |
| amlodipine | 5_tab | a00096 | amlo a2146 | 0.01 | 0.08 |
| amlodipine | 5_tab | a00239 | amlo a0446 | 0.01 | 0.08 |
| amlodipine | 5_tab | a00347 | amlo a0395 | 0.01 | 0.08 |
| amlodipine | 5_tab | a00333 | amlo a00333_inn | 0.01 | 0.08 |
| amlodipine | 5_tab | a00215 | amlo a00215_inn | 0.01 | 0.08 |
| amlodipine | 5_tab | a00367 | amlo a00367_inn | 0.01 | 0.08 |
| amlodipine | 5_tab | a00439 | amlo a00439_inn | 0.00 | 0.08 |
| amlodipine | 5_tab | a00076 | amlo a2991 | 0.00 | 0.16 |
| amlodipine | 5_tab | a00439 | amlo a0268 | 0.00 | 0.08 |
| amlodipine | 5_tab | a00438 | amlo a00438_inn | 0.00 | 0.08 |
| amlodipine | 5_tab | a00232 | amlo a00232_inn | 0.00 | 0.08 |
| amlodipine | 5_tab | a00216 | amlo a00216_inn | 0.00 | 0.16 |
| amlodipine | 5_tab | a00130 | amlo a00130_inn | 0.00 | 0.24 |
| amlodipine | 5_tab | a00130 | amlo a5149 | 0.00 | 0.00 |
| amlodipine | 5_tab | a00004 | amlo a4356 | 0.00 | 0.00 |
| amlodipine | 5_tab | a00192 | amlo a2394 | 0.00 | 0.00 |
| amlodipine | 5_tab | a00280 | amlo a5542 | 0.00 | 0.08 |
| amlodipine | 5_tab | a00334 | amlo a4655 | 0.00 | 0.00 |
| amlodipine | 5_tab | a00082 | amlo a00082_inn | 0.00 | 0.16 |
| amlodipine | 5_tab | a00363 | amlo a00363_inn | 0.00 | 0.08 |
| amlodipine | 5_tab | a00144 | amlo a00144_inn | 0.00 | 0.16 |
| amlodipine | 5_tab | a00311 | amlo a0105 | 0.00 | 0.00 |
| amlodipine | 5_tab | a00149 | amlo a4813 | 0.00 | 0.08 |
| amlodipine | 5_tab | a00387 | amlo a0484 | 0.00 | 0.16 |
| amlodipine | 5_tab | a00287 | amlo a3208 | 0.00 | 0.16 |
| amoxicillin | 125_liq | a00003 | amox a00003_inn | 2.16 | 0.63 |
| amoxicillin | 125_liq | a00215 | amox a2258 | 0.96 | 0.86 |
| amoxicillin | 125_liq | a00283 | amox a2999 | 0.17 | 0.63 |
| amoxicillin | 125_liq | a00309 | amox a00309_inn | 0.15 | 0.16 |
| amoxicillin | 125_liq | a00039 | amox a2891 | 0.14 | 0.31 |
| amoxicillin | 125_liq | a00205 | amox a0065 | 0.06 | 0.24 |
| amoxicillin | 125_liq | a00347 | amox a0413 | 0.06 | 0.08 |
| amoxicillin | 125_liq | a00246 | amox a00246_inn | 0.04 | 0.55 |
| amoxicillin | 125_liq | a00061 | amox a3069 | 0.04 | 0.00 |
| amoxicillin | 125_liq | a00387 | amox a4463 | 0.04 | 0.08 |
| amoxicillin | 125_liq | a00273 | amox a4471 | 0.04 | 0.31 |
| amoxicillin | 125_liq | a00286 | amox a00286_inn | 0.04 | 0.16 |
| amoxicillin | 125_liq | a00082 | amox a3786 | 0.03 | 0.31 |
| amoxicillin | 125_liq | a00309 | amox a0525 | 0.02 | 0.00 |
| amoxicillin | 125_liq | a00061 | amox a00061_inn | 0.00 | 0.08 |
| amoxicillin | 125_liq | a00367 | amox a00367_inn | 0.00 | 0.16 |
| amoxicillin | 125_liq | a00066 | amox a00066_inn | 0.00 | 0.08 |

| | | | | | |
|---|---|---|---|---|---|
| amoxicillin | 125_liq | a00060 | amox a1244 | 0.00 | 0.16 |
| amoxicillin | 125_liq | a00060 | amox a00060_inn | 0.00 | 0.08 |
| amoxicillin | 125_liq | a00243 | amox a4329 | 0.00 | 0.08 |
| amoxicillin | 250_liq | a00003 | amox a00003_inn | 0.57 | 0.08 |
| amoxicillin | 250_liq | a00215 | amox a2258 | 0.16 | 0.08 |
| amoxicillin | 250_liq | a00061 | amox a3069 | 0.01 | 0.08 |
| amoxicillin | 250_liq | a00004 | amox a0601 | 0.00 | 0.08 |
| amoxicillin | 500_cap | a00283 | amox a2999 | 0.85 | 1.73 |
| amoxicillin | 500_cap | a00100 | amox a2789 | 0.04 | 0.16 |
| amoxicillin | 500_cap | a00333 | amox a2650 | 0.04 | 0.24 |
| amoxicillin | 500_cap | a00110 | amox a4355 | 0.00 | 0.08 |
| amoxicillin | 500_tab | a00309 | amox a00309_inn | 15.03 | 2.83 |
| amoxicillin | 500_tab | a00215 | amox a2258 | 0.90 | 0.71 |
| amoxicillin | 500_tab | a00246 | amox a00246_inn | 0.49 | 0.86 |
| amoxicillin | 500_tab | a00286 | amox a00286_inn | 0.38 | 0.31 |
| amoxicillin | 500_tab | a00039 | amox a2891 | 0.38 | 0.24 |
| amoxicillin | 500_tab | a00003 | amox a00003_inn | 0.34 | 0.78 |
| amoxicillin | 500_tab | a00092 | amox a00092_inn | 0.19 | 0.63 |
| amoxicillin | 500_tab | a00246 | amox a4424 | 0.17 | 0.31 |
| amoxicillin | 500_tab | a00061 | amox a3069 | 0.16 | 0.24 |
| amoxicillin | 500_tab | a00309 | amox a0525 | 0.13 | 0.55 |
| amoxicillin | 500_tab | a00387 | amox a4463 | 0.12 | 0.08 |
| amoxicillin | 500_tab | a00367 | amox a4640 | 0.12 | 0.24 |
| amoxicillin | 500_tab | a00205 | amox a0065 | 0.12 | 0.00 |
| amoxicillin | 500_tab | a00004 | amox a0601 | 0.08 | 0.39 |
| amoxicillin | 500_tab | a00076 | amox a2987 | 0.07 | 0.24 |
| amoxicillin | 500_tab | a00412 | amox a2123 | 0.07 | 0.63 |
| amoxicillin | 500_tab | a00334 | amox a2806 | 0.06 | 0.24 |
| amoxicillin | 500_tab | a00116 | amox a0606 | 0.06 | 0.16 |
| amoxicillin | 500_tab | a00082 | amox a3786 | 0.06 | 0.31 |
| amoxicillin | 500_tab | a00347 | amox a0413 | 0.05 | 0.00 |
| amoxicillin | 500_tab | a00021 | amox a3848 | 0.04 | 0.31 |
| amoxicillin | 500_tab | a00367 | amox a00367_inn | 0.04 | 0.00 |
| amoxicillin | 500_tab | a00382 | amox a3611 | 0.04 | 0.00 |
| amoxicillin | 500_tab | a00273 | amox a4471 | 0.03 | 0.31 |
| amoxicillin | 500_tab | a00130 | amox a1680 | 0.03 | 0.24 |
| amoxicillin | 500_tab | a00214 | amox a1332 | 0.01 | 0.31 |
| amoxicillin | 500_tab | a00094 | amox a2461 | 0.01 | 0.00 |
| amoxicillin | 500_tab | a00204 | amox a5379 | 0.01 | 0.00 |
| amoxicillin | 500_tab | a00369 | amox a3551 | 0.01 | 0.31 |
| amoxicillin | 500_tab | a00287 | amox a2238 | 0.01 | 0.00 |
| amoxicillin | 500_tab | a00092 | amox a4709 | 0.00 | 0.31 |
| amoxicillin | 500_tab | a00090 | amox a3633 | 0.00 | 0.24 |
| amoxicillin | 500_tab | a00130 | amox a00130_inn | 0.00 | 0.24 |
| amoxicillin | 500_tab | a00010 | amox a3922 | 0.00 | 0.08 |
| amoxicillin | 500_tab | a00144 | amox a2159 | 0.00 | 0.08 |
| cefixime | 100_cap | a00116 | cefix a00116_inn | 0.73 | 2.83 |
| cefixime | 100_cap | a00309 | cefix a00309_inn | 0.65 | 2.20 |
| cefixime | 100_cap | a00311 | cefix a00311_inn | 0.20 | 0.31 |
| cefixime | 100_cap | a00215 | cefix a4242 | 0.16 | 0.78 |
| cefixime | 100_cap | a00102 | cefix a4132 | 0.08 | 0.94 |
| cefixime | 100_cap | a00412 | cefix a3190 | 0.05 | 0.55 |

| | | | | | |
|---|---|---|---|---|---|
| cefixime | 100_cap | a00423 | cefix a00423_inn | 0.04 | 0.16 |
| cefixime | 100_cap | a00021 | cefix a1893 | 0.03 | 0.55 |
| cefixime | 100_cap | a00333 | cefix a3046 | 0.02 | 0.24 |
| cefixime | 100_cap | a00092 | cefix a00092_inn | 0.02 | 0.55 |
| cefixime | 100_cap | a00113 | cefix a2058 | 0.02 | 0.31 |
| cefixime | 100_cap | a00100 | cefix a4591 | 0.02 | 0.16 |
| cefixime | 100_cap | a00258 | cefix a4910 | 0.01 | 0.24 |
| cefixime | 100_cap | a00116 | cefix a2696 | 0.01 | 0.08 |
| cefixime | 100_cap | a00423 | cefix a1082 | 0.01 | 0.16 |
| cefixime | 100_cap | a00215 | cefix a00215_inn | 0.01 | 0.08 |
| cefixime | 100_cap | a00328 | cefix a4849 | 0.01 | 0.39 |
| cefixime | 100_cap | a00096 | cefix a1473 | 0.01 | 0.08 |
| cefixime | 100_cap | a00009 | cefix a4809 | 0.00 | 0.08 |
| cefixime | 100_cap | a00285 | cefix a00285_inn | 0.00 | 0.16 |
| cefixime | 100_cap | a00438 | cefix a5089 | 0.00 | 0.08 |
| cefixime | 100_cap | a00090 | cefix a0399 | 0.00 | 0.08 |
| cefixime | 100_cap | a00076 | cefix a0501 | 0.00 | 0.00 |
| cefixime | 100_cap | a00439 | cefix a4845 | 0.00 | 0.00 |
| cefixime | 100_cap | a00057 | cefix a1160 | 0.00 | 0.08 |
| cefixime | 100_cap | a00092 | cefix a1174 | 0.00 | 0.24 |
| cefixime | 100_cap | a00214 | cefix a1123 | 0.00 | 0.00 |
| cefixime | 100_cap | a00280 | cefix a4715 | 0.00 | 0.00 |
| cefixime | 200_cap | a00116 | cefix a00116_inn | 0.60 | 0.39 |
| cefixime | 200_cap | a00311 | cefix a00311_inn | 0.23 | 0.16 |
| cefixime | 200_cap | a00438 | cefix a5089 | 0.04 | 0.00 |
| cefixime | 200_cap | a00258 | cefix a4910 | 0.02 | 0.08 |
| cefixime | 200_cap | a00113 | cefix a00113_inn | 0.00 | 0.16 |
| cefixime | 200_tab | a00286 | cefix a00286_inn | 0.07 | 0.00 |
| cefixime | 200_tab | a00309 | cefix a00309_inn | 0.06 | 0.39 |
| cefixime | 200_tab | a00021 | cefix a1893 | 0.06 | 0.08 |
| cefixime | 200_tab | a00328 | cefix a4849 | 0.02 | 0.08 |
| cefixime | 200_tab | a00119 | cefix a4344 | 0.01 | 0.16 |
| cefixime | 200_tab | a00277 | cefix a00277_inn | 0.00 | 0.00 |
| cefixime | 200_tab | a00066 | cefix a00066_inn | 0.00 | 0.00 |
| cefixime | 200_tab | a00277 | cefix a1459 | 0.00 | 0.00 |
| dexamethasone | 0.5_tab | a00234 | dexa a00234_inn | 6.26 | 0.08 |
| dexamethasone | 0.5_tab | a00340 | dexa a4782 | 5.75 | 2.67 |
| dexamethasone | 0.5_tab | a00068 | dexa a5066 | 1.78 | 0.71 |
| dexamethasone | 0.5_tab | a00102 | dexa a5875 | 1.15 | 1.41 |
| dexamethasone | 0.5_tab | a00061 | dexa a5140 | 0.86 | 1.10 |
| dexamethasone | 0.5_tab | a00153 | dexa a0192 | 0.82 | 0.08 |
| dexamethasone | 0.5_tab | a00234 | dexa a5189 | 0.78 | 0.24 |
| dexamethasone | 0.5_tab | a00096 | dexa a1320 | 0.77 | 0.47 |
| dexamethasone | 0.5_tab | a00309 | dexa a4877 | 0.72 | 0.78 |
| dexamethasone | 0.5_tab | a00021 | dexa a3196 | 0.65 | 0.94 |
| dexamethasone | 0.5_tab | a00110 | dexa a00110_inn | 0.61 | 0.39 |
| dexamethasone | 0.5_tab | a00214 | dexa a00214_inn | 0.54 | 0.55 |
| dexamethasone | 0.5_tab | a00287 | dexa a1448 | 0.39 | 0.31 |
| dexamethasone | 0.5_tab | a00246 | dexa a1303 | 0.35 | 0.39 |
| dexamethasone | 0.5_tab | a00149 | dexa a0215 | 0.34 | 0.55 |
| dexamethasone | 0.5_tab | a00110 | dexa a4144 | 0.27 | 0.55 |
| dexamethasone | 0.5_tab | a00333 | dexa a0431 | 0.26 | 1.18 |

| | | | | | |
|---|---|---|---|---|---|
| dexamethasone | 0.5_tab | a00286 | dexa a00286_inn | 0.24 | 0.31 |
| dexamethasone | 0.5_tab | a00246 | dexa a00246_inn | 0.19 | 0.63 |
| dexamethasone | 0.5_tab | a00439 | dexa a3111 | 0.18 | 0.39 |
| dexamethasone | 0.5_tab | a00243 | dexa a00243_inn | 0.16 | 0.16 |
| dexamethasone | 0.5_tab | a00215 | dexa a1635 | 0.10 | 0.00 |
| dexamethasone | 0.5_tab | a00405 | dexa a00405_inn | 0.10 | 0.31 |
| dexamethasone | 0.5_tab | a00367 | dexa a0865 | 0.08 | 0.08 |
| dexamethasone | 0.5_tab | a00100 | dexa a3730 | 0.05 | 0.08 |
| dexamethasone | 0.5_tab | a00094 | dexa a0789 | 0.04 | 0.16 |
| dexamethasone | 0.5_tab | a00385 | dexa a0160 | 0.04 | 0.08 |
| dexamethasone | 0.5_tab | a00161 | dexa a4154 | 0.03 | 0.00 |
| dexamethasone | 0.5_tab | a00010 | dexa a00010_inn | 0.02 | 0.16 |
| dexamethasone | 0.5_tab | a00082 | dexa a2507 | 0.01 | 0.16 |
| dexamethasone | 0.5_tab | a00010 | dexa a0275 | 0.01 | 0.08 |
| dexamethasone | 0.5_tab | a00243 | dexa a3294 | 0.00 | 0.16 |
| dexamethasone | 0.5_tab | a00405 | dexa a2527 | 0.00 | 0.08 |
| dexamethasone | 0.5_tab | a00216 | dexa a00216_inn | 0.00 | 0.08 |
| dexamethasone | 0.5_tab | a00130 | dexa a00130_inn | 0.00 | 0.08 |
| dexamethasone | 0.75_tab | a00340 | dexa a4782 | 2.61 | 0.31 |
| dexamethasone | 0.75_tab | a00110 | dexa a4144 | 1.16 | 0.24 |
| dexamethasone | 0.75_tab | a00061 | dexa a5140 | 0.79 | 0.00 |
| dexamethasone | 0.75_tab | a00116 | dexa a3029 | 0.24 | 0.16 |
| dexamethasone | 0.75_tab | a00294 | dexa a4135 | 0.19 | 0.00 |
| dexamethasone | 0.75_tab | a00215 | dexa a1635 | 0.07 | 0.08 |
| dexamethasone | 0.75_tab | a00287 | dexa a1448 | 0.00 | 0.08 |
| dexamethasone | 0.75_tab | a00243 | dexa a3294 | 0.00 | 0.08 |

| ces, including pharmacies | Samples from unregulated internet vendors | | | Samples from physical sources, including online sales by verified pharmacies | | |
|---|---|---|---|---|---|---|
| Sample weight | Market share | Share of STARmeds samples | Sample weight | Share of market for specific API* | Share of study samples of API | Weight for adjustment within API |
| 1.84 | 0.0000 | 0.47 | 0.0000 | 46.16 | 11.11 | 4.15 |
| 0.42 | 0.0001 | 0.16 | 0.0004 | 6.13 | 6.40 | 0.96 |
| 0.92 | 0.0000 | 0.16 | 0.0000 | 2.81 | 1.35 | 2.09 |
| 0.17 | 0.0000 | 0.08 | 0.0003 | 2.65 | 7.07 | 0.37 |
| 0.43 | 0.0000 | 0.31 | 0.0000 | 2.61 | 2.69 | 0.97 |
| 0.09 | 0.0000 | 0.00 | 0.0000 | 1.57 | 7.74 | 0.20 |
| 0.48 | 0.0000 | 0.00 | 0.0000 | 1.45 | 1.35 | 1.08 |
| 0.19 | 0.0000 | 0.00 | 0.0000 | 1.03 | 2.36 | 0.44 |
| 0.07 | 0.0000 | 0.08 | 0.0001 | 0.77 | 5.05 | 0.15 |
| 0.07 | 0.0000 | 0.16 | 0.0000 | 0.34 | 2.02 | 0.17 |
| 0.07 | 0.0000 | 0.00 | 0.0000 | 0.31 | 2.02 | 0.15 |
| 0.07 | 0.0000 | 0.00 | 0.0000 | 0.25 | 1.68 | 0.15 |
| 0.11 | 0.0000 | 0.08 | 0.0000 | 0.17 | 0.67 | 0.25 |
| 0.16 | 0.0000 | 0.00 | 0.0000 | 0.12 | 0.34 | 0.35 |
| 0.03 | 0.0000 | 0.08 | 0.0000 | 0.10 | 1.35 | 0.08 |
| 0.03 | 0.0000 | 0.00 | 0.0000 | 0.10 | 1.35 | 0.07 |
| 0.01 | 0.0000 | 0.08 | 0.0000 | 0.09 | 2.69 | 0.03 |
| 0.06 | 0.0000 | 0.00 | 0.0000 | 0.09 | 0.67 | 0.13 |
| 0.11 | 0.0000 | 0.00 | 0.0000 | 0.09 | 0.34 | 0.25 |
| 0.01 | 0.0000 | 0.00 | 0.0000 | 0.05 | 1.68 | 0.03 |
| 0.05 | 0.0000 | 0.00 | 0.0000 | 0.04 | 0.34 | 0.12 |
| 0.04 | 0.0000 | 0.00 | 0.0000 | 0.03 | 0.34 | 0.09 |
| 0.01 | 0.0000 | 0.00 | 0.0000 | 0.02 | 1.35 | 0.02 |
| 0.02 | 0.0000 | 0.00 | 0.0000 | 0.02 | 0.67 | 0.03 |
| 0.00 | 0.0000 | 0.08 | 0.0000 | 0.01 | 0.00 | 0.00 |
| 0.02 | 0.0000 | 0.00 | 0.0000 | 0.01 | 0.34 | 0.04 |
| 0.01 | 0.0000 | 0.00 | 0.0000 | 0.01 | 0.34 | 0.03 |
| 0.01 | 0.0000 | 0.00 | 0.0000 | 0.01 | 0.34 | 0.02 |
| 0.00 | 0.0000 | 0.08 | 0.0000 | 0.01 | 0.00 | 0.00 |
| 0.01 | 0.0000 | 0.00 | 0.0000 | 0.01 | 0.34 | 0.02 |
| 0.00 | 0.0000 | 0.00 | 0.0000 | 0.00 | 0.67 | 0.01 |
| 0.00 | 0.0000 | 0.00 | 0.0000 | 0.00 | 0.67 | 0.00 |
| 0.00 | 0.0000 | 0.00 | 0.0000 | 0.00 | 0.34 | 0.00 |
| 0.00 | 0.0000 | 0.08 | 0.0000 | 0.00 | 0.00 | 0.00 |
| 0.00 | 0.0000 | 0.00 | 0.0000 | 0.00 | 0.34 | 0.00 |
| 1.03 | 0.0000 | 0.16 | 0.0001 | 14.15 | 6.06 | 2.33 |
| 0.59 | 0.0000 | 0.16 | 0.0000 | 3.58 | 2.69 | 1.33 |
| 0.80 | 0.0000 | 0.08 | 0.0004 | 3.06 | 1.68 | 1.82 |
| 0.83 | 0.0000 | 0.00 | 0.0000 | 1.90 | 1.01 | 1.88 |
| 0.42 | 0.0000 | 0.00 | 0.0000 | 0.95 | 1.01 | 0.94 |
| 0.29 | 0.0000 | 0.16 | 0.0000 | 0.66 | 1.01 | 0.65 |
| 0.10 | 0.0000 | 0.00 | 0.0000 | 0.32 | 1.35 | 0.23 |
| 0.35 | 0.0000 | 0.00 | 0.0000 | 0.27 | 0.34 | 0.80 |
| 0.12 | 0.0000 | 0.00 | 0.0000 | 0.26 | 1.01 | 0.26 |
| 0.12 | 0.0000 | 0.16 | 0.0000 | 0.19 | 0.67 | 0.28 |
| 0.11 | 0.0000 | 0.00 | 0.0000 | 0.17 | 0.67 | 0.25 |

| | | | | | | |
|---|---|---|---|---|---|---|
| 0.00 | 0.0000 | 0.08 | 0.0000 | 0.12 | 0.00 | 0.00 |
| 0.00 | 0.0000 | 0.08 | 0.0000 | 0.10 | 0.00 | 0.00 |
| 0.00 | 0.0000 | 0.16 | 0.0000 | 0.08 | 0.00 | 0.00 |
| 0.00 | 0.0000 | 0.16 | 0.0000 | 0.07 | 0.00 | 0.00 |
| 0.04 | 0.0000 | 0.00 | 0.0000 | 0.06 | 0.67 | 0.09 |
| 0.00 | 0.0000 | 0.08 | 0.0000 | 0.03 | 0.00 | 0.00 |
| 0.00 | 0.0000 | 0.16 | 0.0000 | 0.03 | 0.00 | 0.00 |
| 0.01 | 0.0000 | 0.00 | 0.0000 | 0.01 | 0.34 | 0.03 |
| 0.00 | 0.0000 | 0.00 | 0.0000 | 0.00 | 0.67 | 0.00 |
| 0.00 | 0.0000 | 0.00 | 0.0000 | 0.00 | 0.34 | 0.01 |
| 0.00 | 0.0000 | 0.08 | 0.0000 | 0.00 | 0.00 | 0.00 |
| 25.03 | 0.0000 | 0.00 | 0.0000 | 19.17 | 1.23 | 15.59 |
| 16.26 | 0.0000 | 0.08 | 0.0001 | 8.30 | 0.82 | 10.13 |
| 23.79 | 0.0000 | 0.00 | 0.0000 | 6.07 | 0.41 | 14.82 |
| 8.52 | 0.0000 | 0.00 | 0.0000 | 4.35 | 0.82 | 5.31 |
| 13.99 | 0.0000 | 0.00 | 0.0000 | 3.57 | 0.41 | 8.72 |
| 6.28 | 0.0000 | 0.00 | 0.0000 | 3.21 | 0.82 | 3.91 |
| 8.12 | 0.0000 | 0.00 | 0.0000 | 2.07 | 0.41 | 5.06 |
| 6.32 | 0.0000 | 0.08 | 0.0000 | 1.61 | 0.41 | 3.93 |
| 1.30 | 0.0000 | 0.00 | 0.0000 | 0.33 | 0.41 | 0.81 |
| 0.36 | 0.0000 | 0.08 | 0.0001 | 0.28 | 1.23 | 0.23 |
| 0.91 | 0.0000 | 0.00 | 0.0000 | 0.23 | 0.41 | 0.57 |
| 0.00 | 0.0000 | 0.08 | 0.0001 | 0.23 | 0.00 | 0.00 |
| 0.73 | 0.0000 | 0.00 | 0.0000 | 0.19 | 0.41 | 0.45 |
| 0.44 | 0.0000 | 0.00 | 0.0000 | 0.11 | 0.41 | 0.27 |
| 0.00 | 0.0000 | 0.16 | 0.0000 | 0.09 | 0.00 | 0.00 |
| 0.25 | 0.0000 | 0.00 | 0.0000 | 0.06 | 0.41 | 0.16 |
| 0.00 | 0.0000 | 0.08 | 0.0000 | 0.06 | 0.00 | 0.00 |
| 0.00 | 0.0000 | 0.08 | 0.0000 | 0.02 | 0.00 | 0.00 |
| 0.00 | 0.0000 | 0.08 | 0.0000 | 0.01 | 0.00 | 0.00 |
| 0.00 | 0.0000 | 0.08 | 0.0000 | 0.00 | 0.00 | 0.00 |
| 0.00 | 0.0000 | 0.00 | 0.0000 | 0.00 | 0.41 | 0.00 |
| 2.31 | 0.0000 | 0.16 | 0.0001 | 10.03 | 6.97 | 1.44 |
| 2.02 | 0.0000 | 0.00 | 0.0000 | 9.30 | 7.38 | 1.26 |
| 1.07 | 0.0000 | 0.00 | 0.0000 | 7.66 | 11.48 | 0.67 |
| 4.07 | 0.0000 | 0.00 | 0.0000 | 5.20 | 2.05 | 2.54 |
| 2.05 | 0.0000 | 0.16 | 0.0001 | 3.14 | 2.46 | 1.28 |
| 1.81 | 0.0000 | 0.08 | 0.0000 | 2.31 | 2.05 | 1.13 |
| 1.97 | 0.0000 | 0.08 | 0.0000 | 1.51 | 1.23 | 1.23 |
| 0.39 | 0.0000 | 0.16 | 0.0002 | 1.19 | 4.92 | 0.24 |
| 0.20 | 0.0000 | 0.16 | 0.0001 | 0.65 | 5.33 | 0.12 |
| 0.36 | 0.0000 | 0.00 | 0.0000 | 0.55 | 2.46 | 0.22 |
| 1.52 | 0.0000 | 0.00 | 0.0000 | 0.39 | 0.41 | 0.95 |
| 0.33 | 0.0000 | 0.08 | 0.0001 | 0.25 | 1.23 | 0.20 |
| 0.32 | 0.0000 | 0.00 | 0.0000 | 0.24 | 1.23 | 0.20 |
| 0.86 | 0.0000 | 0.00 | 0.0000 | 0.22 | 0.41 | 0.53 |
| 0.36 | 0.0000 | 0.00 | 0.0000 | 0.18 | 0.82 | 0.22 |
| 0.69 | 0.0000 | 0.00 | 0.0000 | 0.18 | 0.41 | 0.43 |
| 0.08 | 0.0000 | 0.08 | 0.0001 | 0.17 | 3.28 | 0.05 |
| 0.08 | 0.0000 | 0.00 | 0.0000 | 0.16 | 3.28 | 0.05 |
| 0.27 | 0.0000 | 0.00 | 0.0000 | 0.14 | 0.82 | 0.17 |
| 0.52 | 0.0000 | 0.00 | 0.0000 | 0.13 | 0.41 | 0.32 |

| | | | | | | |
|---|---|---|---|---|---|---|
| 0.16 | 0.0000 | 0.00 | 0.0000 | 0.12 | 1.23 | 0.10 |
| 0.11 | 0.0000 | 0.00 | 0.0000 | 0.11 | 1.64 | 0.07 |
| 0.06 | 0.0000 | 0.00 | 0.0000 | 0.09 | 2.46 | 0.04 |
| 0.33 | 0.0000 | 0.00 | 0.0000 | 0.08 | 0.41 | 0.21 |
| 0.32 | 0.0000 | 0.08 | 0.0000 | 0.08 | 0.41 | 0.20 |
| 0.15 | 0.0000 | 0.00 | 0.0000 | 0.08 | 0.82 | 0.09 |
| 0.28 | 0.0000 | 0.00 | 0.0000 | 0.07 | 0.41 | 0.17 |
| 0.26 | 0.0000 | 0.00 | 0.0000 | 0.07 | 0.41 | 0.16 |
| 0.17 | 0.0000 | 0.08 | 0.0000 | 0.04 | 0.41 | 0.11 |
| 0.00 | 0.0000 | 0.16 | 0.0000 | 0.04 | 0.00 | 0.00 |
| 0.11 | 0.0000 | 0.00 | 0.0000 | 0.03 | 0.41 | 0.07 |
| 0.11 | 0.0000 | 0.00 | 0.0000 | 0.03 | 0.41 | 0.07 |
| 0.10 | 0.0000 | 0.00 | 0.0000 | 0.03 | 0.41 | 0.06 |
| 0.08 | 0.0000 | 0.00 | 0.0000 | 0.02 | 0.41 | 0.05 |
| 0.07 | 0.0000 | 0.00 | 0.0000 | 0.02 | 0.41 | 0.04 |
| 0.07 | 0.0000 | 0.00 | 0.0000 | 0.02 | 0.41 | 0.04 |
| 0.06 | 0.0000 | 0.00 | 0.0000 | 0.02 | 0.41 | 0.04 |
| 0.03 | 0.0000 | 0.00 | 0.0000 | 0.01 | 0.82 | 0.02 |
| 0.05 | 0.0000 | 0.00 | 0.0000 | 0.01 | 0.41 | 0.03 |
| 0.03 | 0.0000 | 0.00 | 0.0000 | 0.01 | 0.41 | 0.02 |
| 0.03 | 0.0000 | 0.00 | 0.0000 | 0.01 | 0.41 | 0.02 |
| 0.02 | 0.0000 | 0.00 | 0.0000 | 0.01 | 0.82 | 0.01 |
| 0.01 | 0.0000 | 0.00 | 0.0000 | 0.01 | 1.23 | 0.01 |
| 0.00 | 0.0000 | 0.08 | 0.0000 | 0.01 | 0.00 | 0.00 |
| 0.00 | 0.0000 | 0.08 | 0.0000 | 0.01 | 0.00 | 0.00 |
| 0.00 | 0.0000 | 0.08 | 0.0000 | 0.01 | 0.00 | 0.00 |
| 0.02 | 0.0000 | 0.00 | 0.0000 | 0.00 | 0.41 | 0.01 |
| 0.00 | 0.0000 | 0.08 | 0.0000 | 0.00 | 0.00 | 0.00 |
| 0.01 | 0.0000 | 0.00 | 0.0000 | 0.00 | 0.82 | 0.00 |
| 0.01 | 0.0000 | 0.00 | 0.0000 | 0.00 | 0.41 | 0.01 |
| 0.00 | 0.0000 | 0.00 | 0.0000 | 0.00 | 0.82 | 0.00 |
| 0.00 | 0.0000 | 0.08 | 0.0000 | 0.00 | 0.00 | 0.00 |
| 0.01 | 0.0000 | 0.00 | 0.0000 | 0.00 | 0.41 | 0.00 |
| 0.00 | 0.0000 | 0.00 | 0.0000 | 0.00 | 0.82 | 0.00 |
| 0.00 | 0.0000 | 0.00 | 0.0000 | 0.00 | 0.82 | 0.00 |
| 3.43 | 0.0000 | 0.00 | 0.0000 | 8.26 | 2.68 | 3.09 |
| 1.12 | 0.0001 | 0.00 | 0.0000 | 3.69 | 3.68 | 1.00 |
| 0.27 | 0.0000 | 0.08 | 0.0002 | 0.65 | 2.68 | 0.24 |
| 0.96 | 0.0000 | 0.08 | 0.0000 | 0.58 | 0.67 | 0.86 |
| 0.45 | 0.0000 | 0.00 | 0.0000 | 0.54 | 1.34 | 0.40 |
| 0.25 | 0.0000 | 0.16 | 0.0000 | 0.23 | 1.00 | 0.22 |
| 0.71 | 0.0000 | 0.00 | 0.0000 | 0.21 | 0.33 | 0.64 |
| 0.08 | 0.0000 | 0.00 | 0.0000 | 0.17 | 2.34 | 0.07 |
| 0.00 | 0.0000 | 0.08 | 0.0001 | 0.17 | 0.00 | 0.00 |
| 0.54 | 0.0000 | 0.08 | 0.0001 | 0.16 | 0.33 | 0.48 |
| 0.12 | 0.0000 | 0.08 | 0.0000 | 0.15 | 1.34 | 0.11 |
| 0.24 | 0.0000 | 0.00 | 0.0000 | 0.15 | 0.67 | 0.22 |
| 0.10 | 0.0000 | 0.00 | 0.0000 | 0.12 | 1.34 | 0.09 |
| 0.00 | 0.0000 | 0.08 | 0.0000 | 0.09 | 0.00 | 0.00 |
| 0.02 | 0.0000 | 0.00 | 0.0000 | 0.01 | 0.33 | 0.02 |
| 0.01 | 0.0000 | 0.00 | 0.0000 | 0.00 | 0.67 | 0.01 |
| 0.01 | 0.0000 | 0.00 | 0.0000 | 0.00 | 0.33 | 0.01 |

| | | | | | | |
|---|---|---|---|---|---|---|
| 0.01 | 0.0000 | 0.00 | 0.0000 | 0.00 | 0.67 | 0.01 |
| 0.01 | 0.0000 | 0.00 | 0.0000 | 0.00 | 0.33 | 0.01 |
| 0.00 | 0.0000 | 0.00 | 0.0000 | 0.00 | 0.33 | 0.00 |
| 7.25 | 0.0000 | 0.00 | 0.0000 | 2.18 | 0.33 | 6.52 |
| 2.02 | 0.0000 | 0.00 | 0.0000 | 0.61 | 0.33 | 1.82 |
| 0.08 | 0.0000 | 0.00 | 0.0000 | 0.03 | 0.33 | 0.08 |
| 0.02 | 0.0000 | 0.00 | 0.0000 | 0.00 | 0.33 | 0.01 |
| 0.49 | 0.0001 | 0.47 | 0.0002 | 3.27 | 7.36 | 0.44 |
| 0.27 | 0.0000 | 0.08 | 0.0001 | 0.17 | 0.67 | 0.25 |
| 0.17 | 0.0000 | 0.08 | 0.0000 | 0.15 | 1.00 | 0.15 |
| 0.02 | 0.0000 | 0.00 | 0.0000 | 0.01 | 0.33 | 0.02 |
| 5.32 | 0.0000 | 0.16 | 0.0003 | 57.59 | 12.04 | 4.78 |
| 1.27 | 0.0001 | 0.00 | 0.0000 | 3.45 | 3.01 | 1.15 |
| 0.56 | 0.0000 | 0.31 | 0.0000 | 1.86 | 3.68 | 0.51 |
| 1.22 | 0.0000 | 0.00 | 0.0000 | 1.47 | 1.34 | 1.10 |
| 1.60 | 0.0000 | 0.08 | 0.0005 | 1.44 | 1.00 | 1.43 |
| 0.43 | 0.0000 | 0.16 | 0.0000 | 1.29 | 3.34 | 0.39 |
| 0.30 | 0.0000 | 0.00 | 0.0000 | 0.72 | 2.68 | 0.27 |
| 0.54 | 0.0000 | 0.24 | 0.0001 | 0.64 | 1.34 | 0.48 |
| 0.68 | 0.0000 | 0.00 | 0.0000 | 0.61 | 1.00 | 0.61 |
| 0.23 | 0.0000 | 0.08 | 0.0002 | 0.48 | 2.34 | 0.20 |
| 1.55 | 0.0000 | 0.08 | 0.0002 | 0.47 | 0.33 | 1.39 |
| 0.50 | 0.0000 | 0.16 | 0.0001 | 0.45 | 1.00 | 0.45 |
| 0.00 | 0.0000 | 0.08 | 0.0001 | 0.44 | 0.00 | 0.00 |
| 0.21 | 0.0000 | 0.08 | 0.0001 | 0.32 | 1.67 | 0.19 |
| 0.29 | 0.0000 | 0.08 | 0.0001 | 0.26 | 1.00 | 0.26 |
| 0.11 | 0.0000 | 0.08 | 0.0001 | 0.26 | 2.68 | 0.10 |
| 0.27 | 0.0000 | 0.08 | 0.0001 | 0.24 | 1.00 | 0.24 |
| 0.40 | 0.0000 | 0.00 | 0.0000 | 0.24 | 0.67 | 0.36 |
| 0.19 | 0.0000 | 0.00 | 0.0000 | 0.23 | 1.34 | 0.17 |
| 0.00 | 0.0000 | 0.08 | 0.0001 | 0.20 | 0.00 | 0.00 |
| 0.14 | 0.0000 | 0.08 | 0.0001 | 0.16 | 1.34 | 0.12 |
| 0.00 | 0.0000 | 0.08 | 0.0000 | 0.15 | 0.00 | 0.00 |
| 0.00 | 0.0000 | 0.08 | 0.0000 | 0.14 | 0.00 | 0.00 |
| 0.09 | 0.0000 | 0.00 | 0.0000 | 0.11 | 1.34 | 0.09 |
| 0.12 | 0.0000 | 0.00 | 0.0000 | 0.10 | 1.00 | 0.10 |
| 0.05 | 0.0000 | 0.00 | 0.0000 | 0.06 | 1.34 | 0.04 |
| 0.00 | 0.0000 | 0.08 | 0.0000 | 0.05 | 0.00 | 0.00 |
| 0.00 | 0.0000 | 0.16 | 0.0000 | 0.03 | 0.00 | 0.00 |
| 0.02 | 0.0000 | 0.00 | 0.0000 | 0.03 | 1.34 | 0.02 |
| 0.00 | 0.0000 | 0.08 | 0.0000 | 0.02 | 0.00 | 0.00 |
| 0.01 | 0.0000 | 0.00 | 0.0000 | 0.02 | 1.34 | 0.01 |
| 0.01 | 0.0000 | 0.00 | 0.0000 | 0.01 | 1.00 | 0.01 |
| 0.01 | 0.0000 | 0.16 | 0.0000 | 0.01 | 1.00 | 0.01 |
| 0.02 | 0.0000 | 0.16 | 0.0000 | 0.01 | 0.33 | 0.02 |
| 0.01 | 0.0000 | 0.00 | 0.0000 | 0.00 | 0.33 | 0.01 |
| 0.26 | 0.0000 | 0.55 | 0.0000 | 19.41 | 18.46 | 1.05 |
| 0.30 | 0.0000 | 0.16 | 0.0000 | 17.42 | 14.36 | 1.21 |
| 0.65 | 0.0000 | 0.08 | 0.0000 | 5.44 | 2.05 | 2.65 |
| 0.21 | 0.0000 | 0.24 | 0.0001 | 4.30 | 5.13 | 0.84 |
| 0.09 | 0.0000 | 0.00 | 0.0000 | 2.15 | 6.15 | 0.35 |
| 0.09 | 0.0000 | 0.08 | 0.0000 | 1.34 | 3.59 | 0.37 |

| | | | | | | |
|---|---|---|---|---|---|---|
| 0.23 | 0.0000 | 0.00 | 0.0000 | 0.97 | 1.03 | 0.94 |
| 0.05 | 0.0000 | 0.00 | 0.0000 | 0.79 | 3.59 | 0.22 |
| 0.10 | 0.0000 | 0.08 | 0.0000 | 0.62 | 1.54 | 0.40 |
| 0.04 | 0.0000 | 0.00 | 0.0000 | 0.55 | 3.59 | 0.15 |
| 0.06 | 0.0000 | 0.00 | 0.0000 | 0.50 | 2.05 | 0.24 |
| 0.11 | 0.0000 | 0.00 | 0.0000 | 0.45 | 1.03 | 0.44 |
| 0.05 | 0.0000 | 0.00 | 0.0000 | 0.34 | 1.54 | 0.22 |
| 0.13 | 0.0000 | 0.00 | 0.0000 | 0.27 | 0.51 | 0.52 |
| 0.05 | 0.0000 | 0.08 | 0.0000 | 0.22 | 1.03 | 0.22 |
| 0.10 | 0.0000 | 0.00 | 0.0000 | 0.21 | 0.51 | 0.41 |
| 0.01 | 0.0000 | 0.08 | 0.0000 | 0.16 | 2.56 | 0.06 |
| 0.06 | 0.0000 | 0.08 | 0.0000 | 0.13 | 0.51 | 0.26 |
| 0.05 | 0.0000 | 0.08 | 0.0000 | 0.10 | 0.51 | 0.20 |
| 0.02 | 0.0000 | 0.00 | 0.0000 | 0.10 | 1.03 | 0.10 |
| 0.04 | 0.0000 | 0.00 | 0.0000 | 0.08 | 0.51 | 0.16 |
| 0.03 | 0.0000 | 0.00 | 0.0000 | 0.07 | 0.51 | 0.13 |
| 0.00 | 0.0000 | 0.08 | 0.0000 | 0.03 | 0.00 | 0.00 |
| 0.00 | 0.0000 | 0.08 | 0.0000 | 0.02 | 0.00 | 0.00 |
| 0.01 | 0.0000 | 0.00 | 0.0000 | 0.02 | 0.51 | 0.03 |
| 0.00 | 0.0000 | 0.00 | 0.0000 | 0.02 | 1.54 | 0.01 |
| 0.00 | 0.0000 | 0.16 | 0.0000 | 0.01 | 0.00 | 0.00 |
| 0.00 | 0.0000 | 0.08 | 0.0000 | 0.00 | 0.00 | 0.00 |
| 1.52 | 0.0000 | 0.08 | 0.0000 | 15.94 | 2.56 | 6.22 |
| 1.44 | 0.0000 | 0.00 | 0.0000 | 6.04 | 1.03 | 5.89 |
| 0.00 | 0.0000 | 0.08 | 0.0000 | 1.07 | 0.00 | 0.00 |
| 0.25 | 0.0000 | 0.00 | 0.0000 | 0.52 | 0.51 | 1.01 |
| 0.00 | 0.0000 | 0.00 | 0.0000 | 0.01 | 1.03 | 0.01 |
| 0.00 | 0.0000 | 0.08 | 0.0000 | 1.91 | 0.00 | 0.00 |
| 0.16 | 0.0000 | 0.00 | 0.0000 | 1.63 | 2.56 | 0.63 |
| 0.71 | 0.0000 | 0.00 | 0.0000 | 1.49 | 0.51 | 2.90 |
| 0.31 | 0.0000 | 0.08 | 0.0000 | 0.65 | 0.51 | 1.26 |
| 0.08 | 0.0000 | 0.08 | 0.0000 | 0.35 | 1.03 | 0.34 |
| 0.00 | 0.0000 | 0.08 | 0.0000 | 0.11 | 0.00 | 0.00 |
| 0.00 | 0.0000 | 0.08 | 0.0000 | 0.05 | 0.00 | 0.00 |
| 0.00 | 0.0000 | 0.08 | 0.0000 | 0.01 | 0.00 | 0.00 |
| 79.80 | 0.0000 | 0.00 | 0.0000 | 21.52 | 0.42 | 51.43 |
| 2.15 | 0.0006 | 0.08 | 0.0073 | 19.74 | 14.23 | 1.39 |
| 2.51 | 0.0002 | 0.08 | 0.0023 | 6.10 | 3.77 | 1.62 |
| 0.81 | 0.0001 | 0.24 | 0.0004 | 3.94 | 7.53 | 0.52 |
| 0.78 | 0.0001 | 0.00 | 0.0000 | 2.95 | 5.86 | 0.50 |
| 10.39 | 0.0001 | 0.00 | 0.0000 | 2.80 | 0.42 | 6.69 |
| 3.32 | 0.0001 | 0.16 | 0.0005 | 2.68 | 1.26 | 2.14 |
| 1.63 | 0.0001 | 0.31 | 0.0002 | 2.64 | 2.51 | 1.05 |
| 0.92 | 0.0001 | 0.00 | 0.0000 | 2.48 | 4.18 | 0.59 |
| 0.69 | 0.0001 | 0.24 | 0.0003 | 2.24 | 5.02 | 0.45 |
| 1.57 | 0.0000 | 0.00 | 0.0000 | 2.11 | 2.09 | 1.01 |
| 0.98 | 0.0000 | 0.00 | 0.0000 | 1.85 | 2.93 | 0.63 |
| 1.24 | 0.0000 | 0.08 | 0.0005 | 1.33 | 1.67 | 0.80 |
| 0.89 | 0.0000 | 0.00 | 0.0000 | 1.20 | 2.09 | 0.57 |
| 0.61 | 0.0000 | 0.00 | 0.0000 | 1.16 | 2.93 | 0.39 |
| 0.49 | 0.0000 | 0.00 | 0.0000 | 0.92 | 2.93 | 0.31 |
| 0.22 | 0.0000 | 0.08 | 0.0003 | 0.90 | 6.28 | 0.14 |

| | | | | | | |
|---|---|---|---|---|---|---|
| 0.77 | 0.0000 | 0.00 | 0.0000 | 0.83 | 1.67 | 0.50 |
| 0.30 | 0.0000 | 0.08 | 0.0000 | 0.64 | 3.35 | 0.19 |
| 0.45 | 0.0000 | 0.00 | 0.0000 | 0.60 | 2.09 | 0.29 |
| 1.05 | 0.0000 | 0.00 | 0.0000 | 0.57 | 0.84 | 0.68 |
| 0.00 | 0.0000 | 0.08 | 0.0001 | 0.36 | 0.00 | 0.00 |
| 0.31 | 0.0000 | 0.24 | 0.0000 | 0.33 | 1.67 | 0.20 |
| 1.00 | 0.0000 | 0.00 | 0.0000 | 0.27 | 0.42 | 0.65 |
| 0.64 | 0.0000 | 0.08 | 0.0001 | 0.17 | 0.42 | 0.42 |
| 0.28 | 0.0000 | 0.00 | 0.0000 | 0.15 | 0.84 | 0.18 |
| 0.54 | 0.0000 | 0.00 | 0.0000 | 0.15 | 0.42 | 0.35 |
| 0.00 | 0.0000 | 0.08 | 0.0000 | 0.12 | 0.00 | 0.00 |
| 0.15 | 0.0000 | 0.00 | 0.0000 | 0.08 | 0.84 | 0.09 |
| 0.07 | 0.0000 | 0.00 | 0.0000 | 0.04 | 0.84 | 0.04 |
| 0.07 | 0.0000 | 0.00 | 0.0000 | 0.02 | 0.42 | 0.04 |
| 0.03 | 0.0000 | 0.08 | 0.0000 | 0.02 | 0.84 | 0.02 |
| 0.06 | 0.0000 | 0.00 | 0.0000 | 0.02 | 0.42 | 0.04 |
| 0.04 | 0.0000 | 0.00 | 0.0000 | 0.01 | 0.42 | 0.03 |
| 0.00 | 0.0000 | 0.00 | 0.0000 | 0.00 | 0.42 | 0.00 |
| 8.30 | 0.0003 | 0.00 | 0.0000 | 8.96 | 1.67 | 5.35 |
| 4.94 | 0.0001 | 0.16 | 0.0007 | 4.00 | 1.26 | 3.18 |
| 0.00 | 0.0001 | 0.08 | 0.0010 | 2.72 | 0.00 | 0.00 |
| 1.53 | 0.0000 | 0.08 | 0.0003 | 0.82 | 0.84 | 0.98 |
| 0.00 | 0.0000 | 0.08 | 0.0002 | 0.66 | 0.00 | 0.00 |
| 0.88 | 0.0000 | 0.16 | 0.0000 | 0.24 | 0.42 | 0.57 |
| 0.04 | 0.0000 | 0.00 | 0.0000 | 0.01 | 0.42 | 0.02 |
| 0.02 | 0.0000 | 0.00 | 0.0000 | 0.01 | 0.42 | 0.01 |
